# Standardised evaluation and monitoring of site-specific AI performance with physical CT phantoms

**DOI:** 10.64898/2026.07.01.26357033

**Authors:** Ulrich Genske, Angelo Laudani, Li Yan, Yang Peng, Georg Böning, Sevtap Tugce Ulas, Moritz P. Wagner, Torsten Diekhoff, Bernd Hamm, Paul Jahnke

## Abstract

Clinical deployment of medical imaging artificial intelligence (AI) requires objective and continuous quality assurance, yet standardised methods for this purpose have not been established. Here, we present a framework using physical phantoms for standardised on-site testing and monitoring of AI, demonstrated in CT-based liver lesion detection. We begin by designing phantoms tailored to the anatomical input domain expected by AI algorithms, and then systematically assess how AI performance is affected by variations in scanner technology and operation across two clinical CT systems. Next, we perform longitudinal monitoring, yielding consistent results over fifteen months on both systems. Finally, we validate clinical relevance by demonstrating that AI models trained on phantom data generalize effectively to patients and exhibit no evidence of phantom-specific adaptation. Our findings show that clinically realistic phantoms enable standardised, site-specific testing and monitoring of AI, providing a proactive method for local and cross-institutional quality assurance.

---

The integration of artificial intelligence (AI) into medical imaging workflows is expanding across modalities and clinical applications, with AI systems now supporting tasks ranging from lesion detection to disease classification and workflow triage. However, despite rapid progress in algorithm development and regulatory authorization, concerns about trust, transparency, and reliability remain substantial barriers to broader automation and more effective integration^1,2^.

AI models learn patterns from the characteristics of their training data, so shifts in these characteristics can degrade performance when clinical imaging falls outside the model’s expected input domain^3^. In medical imaging, a major source of such shifts is the inherent heterogeneity of the imaging modalities themselves^2^. Clinical image appearance is not standardised and varies widely due to differences in scanner models, acquisition protocols, and a growing range of reconstruction and post-processing techniques, all of which differ between sites and evolve over time. These variations can alter the image features that AI models depend on, leading to performance degradation when data quality does not align with model expectations^4,5^. Even with regulatory authorization, AI applications can therefore exhibit unexpected site-level performance issues^6,7^. Recognizing these challenges, professional practice parameters outline responsibilities for local acceptance testing and performance monitoring^8^. However, a standardised framework to systematically evaluate and monitor how local imaging conditions affect AI performance has not yet been established.

Current AI monitoring strategies rely largely on tracking performance during clinical deployment, which offers valuable real-world insights^9^ but has important limitations for standardised proactive quality assurance. These limitations arise from the reactive nature of monitoring, which detects problems only after they occur in real patients, making close radiologist oversight necessary to ensure patient safety. They also result from the inherent constraints of testing exclusively in patients, who cannot undergo repeated test scans, are not identical or easy to access, and lack a defined ground truth. As a consequence, proactive or standardised testing is precluded, comparability of assessments is limited, and expert annotations are required, which add work and introduce bias. These limitations make it difficult to systematically monitor imaging and AI systems or investigate specific failure modes across time, sites, and technologies^2,10^.

Physical phantoms have long played a central role in testing and monitoring imaging systems^11,12^, offering a standardised and reproducible method of quality assurance. They serve as consistent imaging targets that can be deployed across institutions and over time. While conventional phantom testing focuses on physical metrics and basic scanner performance, phantoms can also be adapted to evaluate clinical tasks such as lesion detection^13,14^. This enables direct assessment of diagnostic performance in relation to image quality, treating the observer (e.g., a radiologist) as the test subject^15,16^. This same principle can be extended to AI, where the algorithm functions as the observer. However, for phantoms to be effective in AI, phantom imaging data must align with the algorithm’s input requirements by replicating the relevant clinical context, including relevant anatomical and pathological features.

Here, we present a framework that uses such task-matched physical phantoms for standardised prospective testing and monitoring of AI models, demonstrated in CT-based liver lesion detection. We begin by designing anatomically realistic phantoms that replicate the imaging characteristics of hepatic lesions. We then use these phantoms to systematically evaluate AI performance as imaging parameters and lesion characteristics vary across two clinical CT systems. We repeat these assessments over fifteen months and demonstrate reproducibility of our findings. Finally, we validate clinical relevance by showing that AI models trained on phantom data generalize effectively to three independent patient cohorts and exhibit no evidence of phantom-specific adaptation.

## Results

### Anatomical domain-matched phantom design

Imaging phantoms are physical objects designed to enable proactive, controlled scan data collection and testing of imaging systems. Traditional phantoms typically feature simple geometric shapes and are primarily used for physics-based evaluations. In contrast, anatomically realistic phantoms that replicate human anatomy, pathology, and tissue detail offer the potential to extend these benefits to the assessment of clinical imaging and diagnostic performance. Advances in manufacturing technologies now make it possible to fabricate such phantoms directly from medical image data, enabling precise modeling of desired anatomy and pathology^17–20^. Building on these developments, we designed anatomically detailed CT phantoms for evaluating AI-based liver lesion detection.

Two abdominal CT phantoms with synthetically inserted liver lesions were first virtually designed and then physically manufactured (**Fig. 1a**). The design process began by retrospectively selecting two portal venous phase CT scans from patients without any liver lesions. Forty-two spherical lesions were then digitally inserted into each liver. Six lesions were included for each combination of 8 or 12 mm lesion diameters and -10, -20, -30 or -40 HU lesion contrast relative to the surrounding liver tissue, except for the -10 HU group, which contained only 12 mm lesions. The final virtual datasets were fabricated into physical CT phantoms using inkjet and 3D printing technology based on previously established methods^17,18^. This process then resulted in pairs of virtual and physical phantoms, with the two physical phantoms replicating the lesion information defined in their respective virtual designs (**Fig. 1b-d**).

**Fig. 1.**
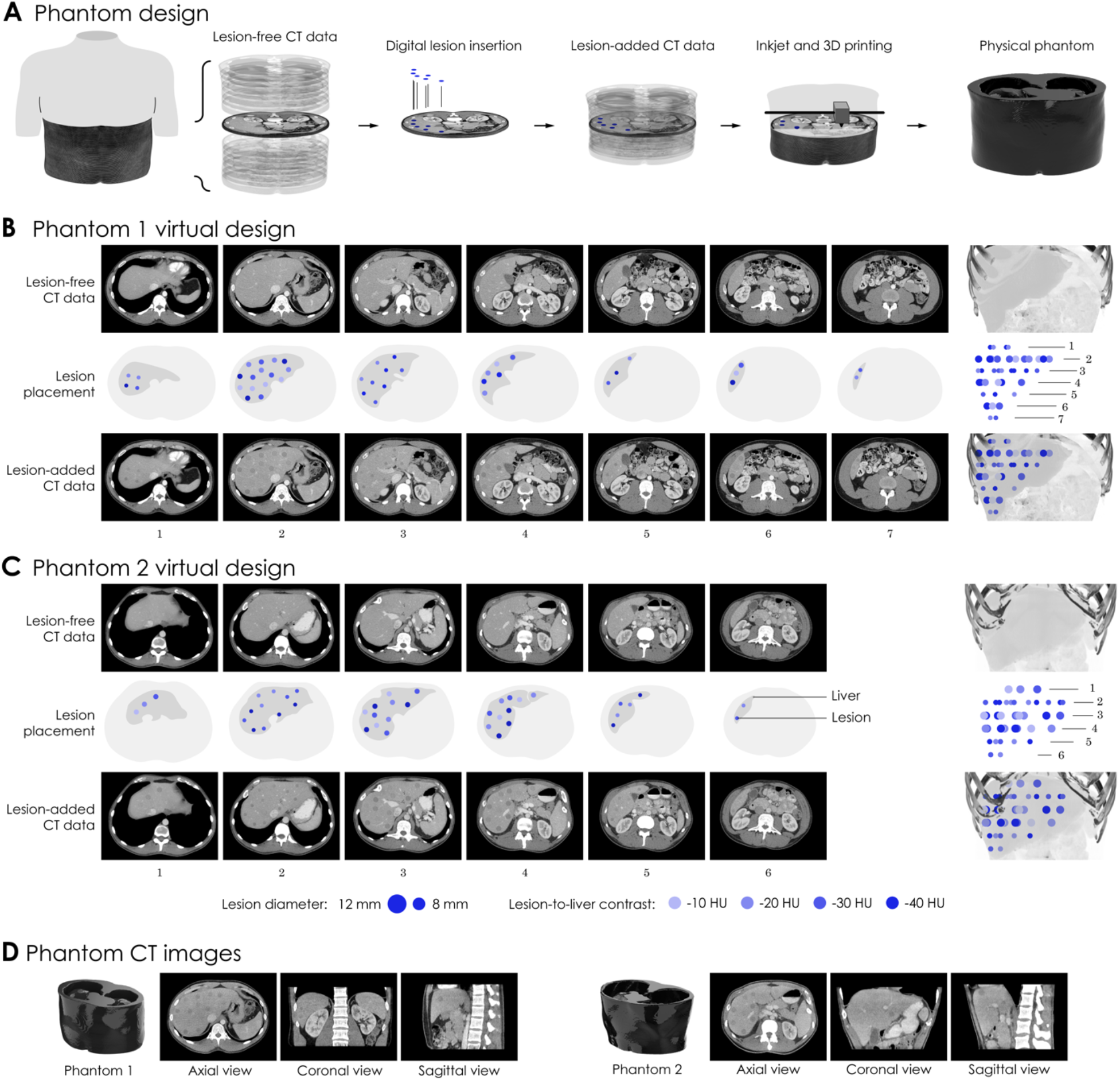
CT phantom design. **a**, Phantom design began with synthetic insertion of spherical liver lesions into abdominal CT data. The data were processed into physical phantoms using inkjet and 3D printing. **b**, Abdominal CT data used for Phantom 1, lesion placement, and resulting CT data with synthetically inserted lesions. Illustrations indicate lesion placement across seven sections. **c**, Abdominal CT data used for Phantom 2, lesion placement, and resulting CT data with synthetically inserted lesions. Illustrations indicate lesion placement across six sections. **d**, Axial, coronal, and sagittal CT images of the phantoms acquired at 10.5 mGy and reconstructed using hybrid iterative reconstruction.

### AI performance testing on clinical CT scanners

The liver is a common site for both primary lesions and metastatic disease^21,22^, and the presence of liver lesions can alter therapeutic strategies and affect patient outcomes. We trained two previously published AI models for liver lesion detection and segmentation^23,24^ using a publicly available patient dataset^25^ and tested their performance on two clinical scanner systems.

The nnU-Net framework provides out-of-the-box deep learning models with state-of-the-art performance and is widely used as a baseline and development platform for medical image detection and segmentation tasks^23^. The Liver Tumor Segmentation (LiTS) challenge was released to advance AI-based liver lesion detection and segmentation and provides a benchmark for model development and comparison^25^. As part of this challenge, 131 annotated patient datasets with 885 liver lesions were made publicly available. We implemented two of the highest-performing nnU-Net configurations using this dataset: the 3D full-resolution architecture in its original version (3D-ov) and an updated variant with residual connections in the encoder path (3D-res) (**Fig. 2a**).

**Fig. 2.**
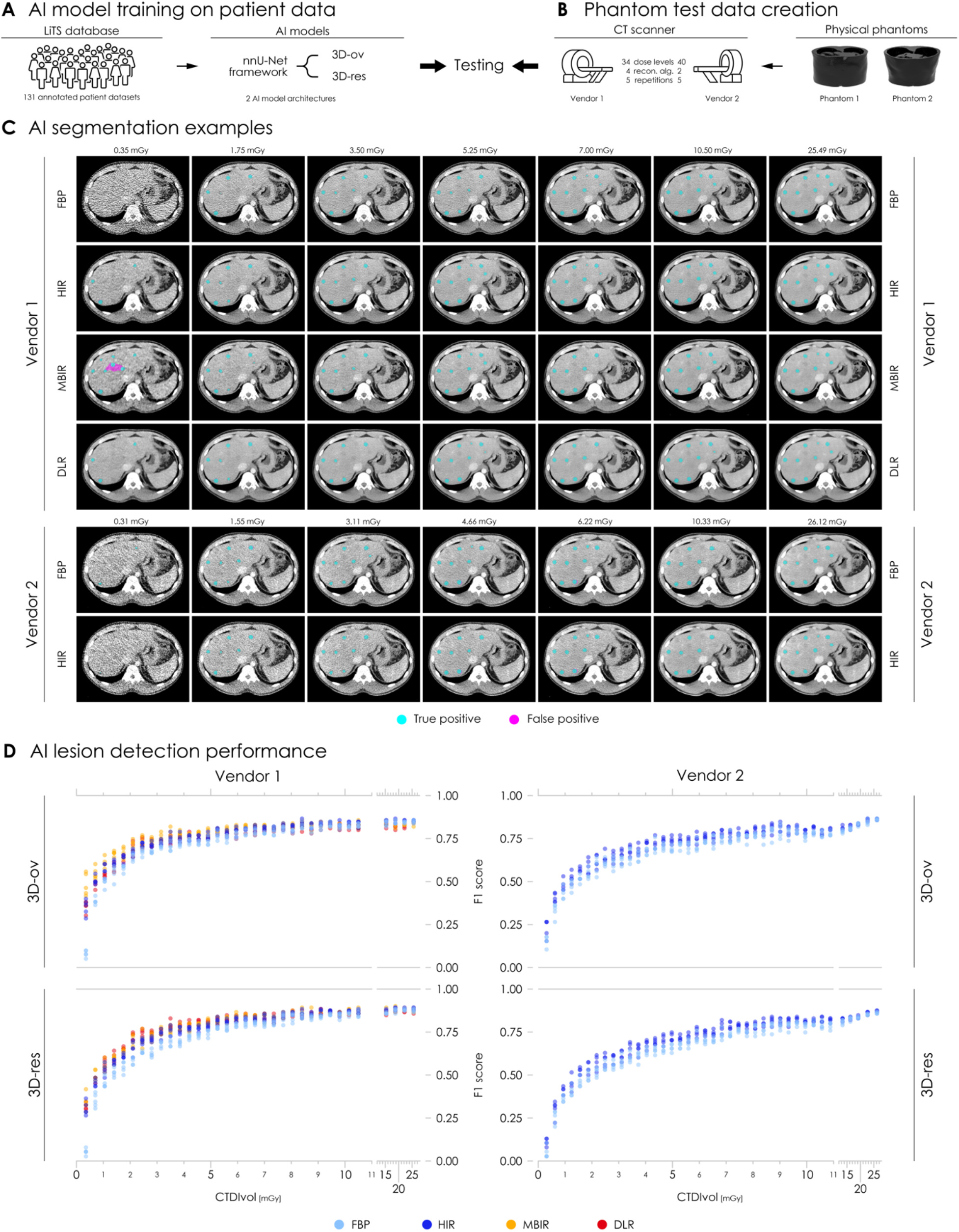
AI liver lesion detection performance across two CT scanners. **a**, Implementation of two AI models trained on the Liver Tumor Segmentation (LiTS) patient dataset using the nnU-Net framework’s original 3D full-resolution architecture (3D-ov) and a modified variant with residual connections in the encoder path (3D-res). **b**, Test data were generated by repeatedly scanning both liver lesion-containing phantoms on two clinical scanners from different vendors, using dose levels from 0.3 to 26.1 mGy and all available image reconstruction algorithms (recon. alg.). **c**, Examples of liver lesion segmentation on CT images from Phantom 1 using the 3D-ov U-Net model across both scanners, varying dose levels, and all available image reconstruction algorithms: filtered back projection (FBP), hybrid iterative reconstruction (HIR), model-based iterative reconstruction (MBIR), and deep-learning reconstruction (DLR). **d**, Liver lesion detection performance of both U-Net models across both scanners, all dose levels, repeated acquisitions, and reconstruction algorithms. CTDIvol is the volume computed tomography dose index.

To evaluate model performance, both phantoms described in the previous section were scanned on two clinical CT systems from different vendors, with systematic variation in radiation dose and image reconstruction (**Fig. 2b**). Scan doses ranged from 0.31 to 26.1 mGy, and all available reconstruction algorithms were used, with five repeated acquisitions per dose-reconstruction combination. Each scanner’s standard clinical oncologic abdominal imaging slice thickness and reconstruction increment were used: 1/0.8 mm for Vendor 1 and 0.625/0.625 mm for Vendor 2. We applied both AI models to the resulting 2,160 scan datasets and evaluated liver lesion detection performance against the virtual ground truth defined during phantom design. Lesion overlap was assessed using Dice scores, and detection outcomes were categorized as true positives (TP), false positives (FP), and false negatives (FN). These metrics were then used to calculate precision (TP/[TP+FP]), recall (TP/[TP+FN]), and the F1 score, defined as the harmonic mean of precision and recall.

**Figure 2c** presents examples of AI-based liver lesion segmentation across varying dose levels, both scanners, and all reconstruction algorithms. F1 scores across all datasets are shown in **Figure 2d**. Lesions with -10 HU contrast were excluded from the analysis as they consistently yielded F1 scores of zero. Both AI models exhibited a dose-dependent decline in lesion detection, with a pronounced decrease at lower dose levels on both scanner systems. Because false positives were rare (median: 0; maximum: 9 at 0.31 mGy), the F1 scores closely reflected recall, while precision remained high across most dose levels (**Supplementary Fig. 1**). Filtered back projection (FBP) yielded lower detection performance than hybrid iterative reconstruction (HIR) across both models and scanners, with mean area under the curve (AUC) differences across all dose levels ranging from 0.032 to 0.045 (**Supplementary Table 2**). Among the denoising iterative and deep learning reconstruction algorithms available from Vendor 1, differences were small and no consistent pattern was observed across model architectures. **Supplementary Figure 2** shows AUC values for all image reconstruction methods, model architectures, and scanners. As both scanners supported HIR, all subsequent analyses were performed using HIR data only.

### Impact of scanner model, AI architecture, and lesion characteristics

Since the same phantoms were scanned on both scanners and evaluated by both AI models, a direct comparison of AI performance across scanner systems and model architectures was possible, in addition to the analysis of dose levels and reconstruction algorithms presented above. Furthermore, because the phantoms were designed to include a controlled range of liver lesion sizes and contrasts, a subanalysis of lesion groups could be conducted to assess how lesion characteristics affect AI detection performance. Comparative analysis between scanners showed that both AI models consistently achieved higher performance on data from Vendor 1. As this scanner system employed a thicker slice thickness compared to Vendor 2 and image slice thickness has been reported to influence AI performance^26^, we investigated whether this difference alone could explain the observed performance gap. To test this, we linearly interpolated the data from Vendor 2 to match the slice thickness used by Vendor 1 and re-evaluated the models on the interpolated datasets. Despite this adjustment, both models continued to perform better on the data from Vendor 1, effectively confirming their superior performance on this scanner system (**Supplementary Table 2**, **Supplementary Fig. 3**).

The original 3D nnU-Net architecture (3D-ov) uses a classic encoder-decoder structure with standard convolutional blocks, while the newer ResEnc L variant (3D-res) incorporates residual connections in the encoder to enhance training stability and segmentation performance. Despite these architectural differences, detection performance was comparable between models, with only marginal differences in F1 scores and AUC values across both scanners (**Supplementary Table 2**, **Supplementary Fig. 4**). The 3D-ov architecture showed slightly superior performance on Vendor 2 data, particularly at lower doses, while no consistent advantage of either architecture was observed on Vendor 1 data.

**Figure 3** presents the F1 score results from the lesion subgroup analysis and shows that the largest and most conspicuous lesions were reliably detected even at lower doses, while detection performance declined as lesion size and contrast decreased. Consistent with this, smaller, lower-contrast lesions required higher radiation doses for detection than larger, higher-contrast lesions. These findings align with expectations and confirm that the phantoms effectively modeled varying degrees of lesion complexity for evaluating AI performance. Per lesion subgroup recall and precision results are provided in **Supplementary Figure 5**.

**Fig. 3.**
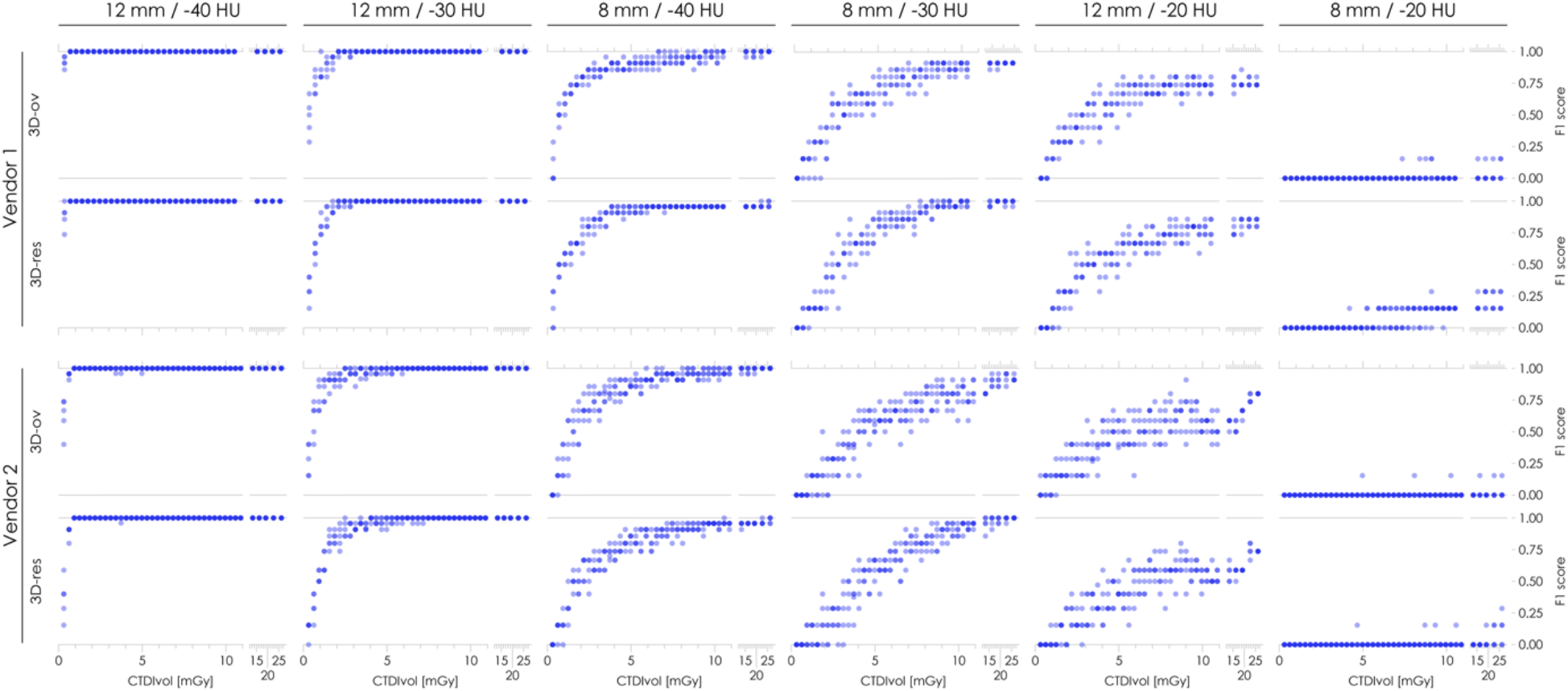
AI detection performance across liver lesion subgroups. Results are presented for each lesion diameter and lesion-to-liver contrast combination labelled at the top of each panel across both CT scanner systems and nnU-Net model architectures: the original 3D full-resolution configuration (3D-ov) and a modified variant with residual connections in the encoder path (3D-res). CTDIvol is the volume computed tomography dose index.

These findings provided insights into the performance characteristics and sensitivity of the two AI models to dose, image reconstruction, lesion characteristics, and differences between scanner systems and model architectures. As the testing protocol was standardised and repeatable, the resulting data also served as a baseline for assessing performance consistency over time in the next phase of our investigation.

### Longitudinal monitoring of AI performance

Local monitoring of AI systems is critical for ensuring reliable and trustworthy AI deployment. However, standardised or proactive testing directly in patients is not feasible due to ethical and practical constraints. In contrast, phantoms provide consistent imaging targets and support repeated testing and direct comparisons of results over time. Using this approach, we performed follow-up testing on both scanner systems fourteen months after initial testing as well as an additional test one month later on each scanner. During this period, none of the scanners underwent major hardware or software updates or mechanical part replacements.

Longitudinal monitoring F1 score results are shown in **Figure 4**. Recall and precision results are provided in **Supplementary Figure 6**. AI detection performance was stable and reproducible during the 15-month monitoring period across both scanner systems and AI architectures. The results thus confirmed system consistency over time. For practical implementation in routine quality assurance workflows, this analysis can be streamlined by focusing on dose levels below 11 mGy and summarizing performance using normalized AUC values ranging from 0 (no detection) to 1 (perfect detection). This approach limits scan time, reduces tube heating, and provides a concise overview suitable for regular monitoring. Bar graphs in **Figure 4** show a condensed version of our monitoring results using this analysis, numerical results are compiled in **Supplementary Table 3**.

**Fig. 4.**
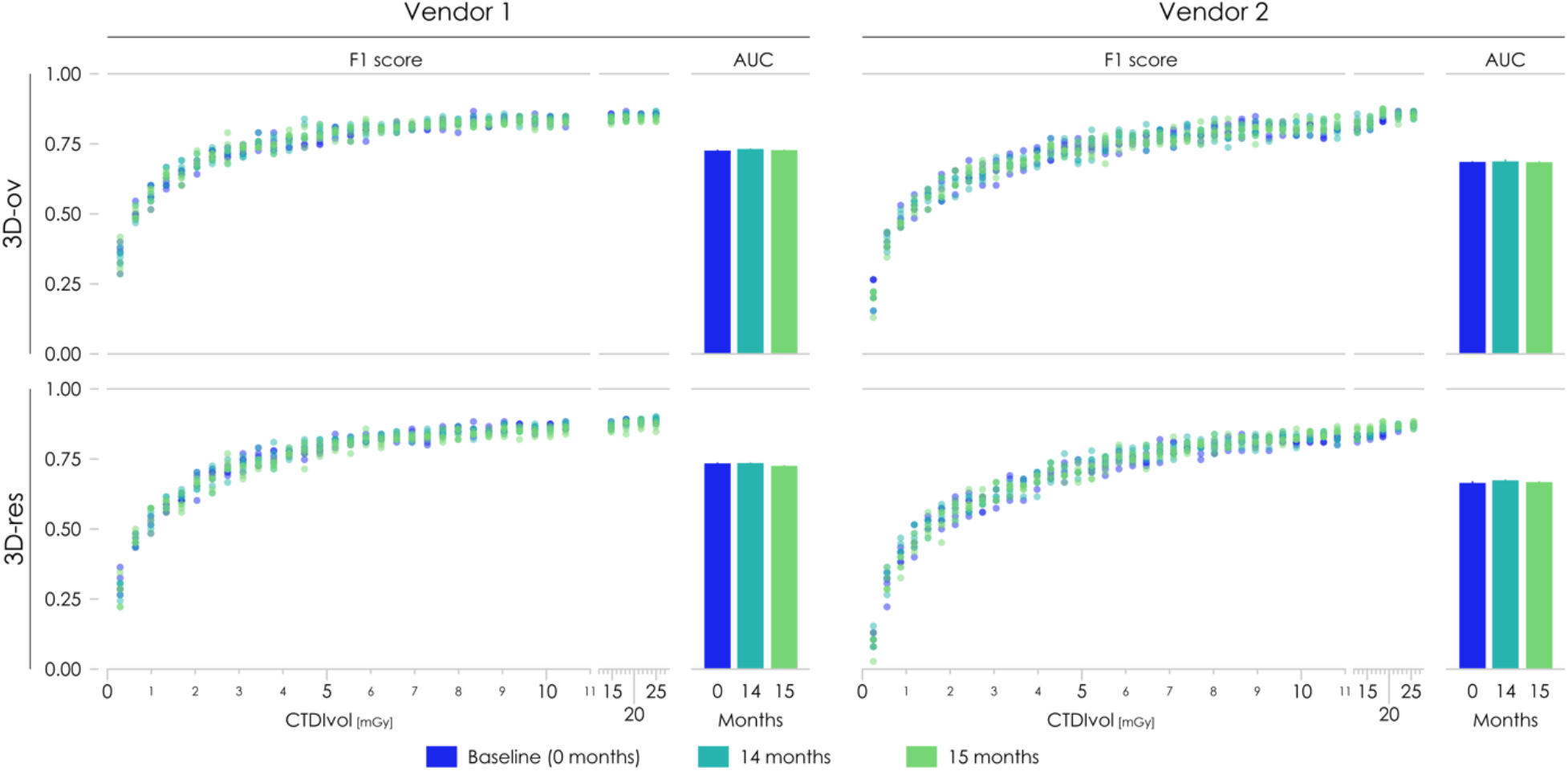
Longitudinal monitoring of AI performance. Liver lesion detection at baseline (0 months) and after repeated acquisitions at 14 and 15 months. Results are presented across both CT scanner systems and nnU-Net model architectures: the original 3D full-resolution configuration (3D-ov) and a modified variant with residual connections in the encoder path (3D-res). CTDIvol is the volume computed tomography dose index. Bar graphs show mean normalised area-under-the-curve (AUC) values across five repeated acquisitions from a subanalysis including only dose levels below 11 mGy. Error bars indicate ranges.

### Generalisability of phantom-trained AI models to clinical data

Phantoms were designed to closely replicate human anatomy and contain liver lesions spanning a range of sizes and contrasts, from smaller, lower-contrast to larger, more conspicuous lesions. AI lesion detection followed expected patterns across these varying levels of lesion conspicuity, suggesting that the phantoms matched the input domain expected by the AI models. To further strengthen this evidence, we tested whether models trained exclusively on phantom-derived scan data could effectively generalise to real clinical patient cohorts. We used a parallel training approach and trained both AI architectures separately on either phantom-only or patient-only data. Model performance was then compared on the same three independent clinical test cohorts.

The phantom-only training set consisted of five scans from five different phantoms: the two phantoms used for testing in the previous sections, a modified version of phantom 1 with altered synthetic lesion design, and two additional phantoms reproducing patients with naturally occurring liver lesions. The corresponding patient-only training set consisted of the patient data used to generate these five phantoms and therefore included the same anatomies and liver lesions (**Fig. 5a**). A second patient-only training set was assembled by selecting five training cases from the LiTS training cohort to match the lesion count of the other training sets (**Fig. 5b**). However, as this patient data could not be proactively designed or manipulated, a perfect match in lesion properties was not achievable. Consequently, the phantom and associated patient data contained smaller and lower-contrast lesions than the LiTS-derived patient dataset (**Supplementary Fig. 7**). Both the 3D-ov and 3D-res nnU-Net configurations were trained on each of the three datasets independently and subsequently evaluated on three independent clinical test cohorts: the remaining scans from the LiTS training cohort (excluding the five cases used for model training), the LiTS test cohort, and an in-house cohort retrospectively collected from our clinical database (**Fig. 5c**).

**Fig. 5.**
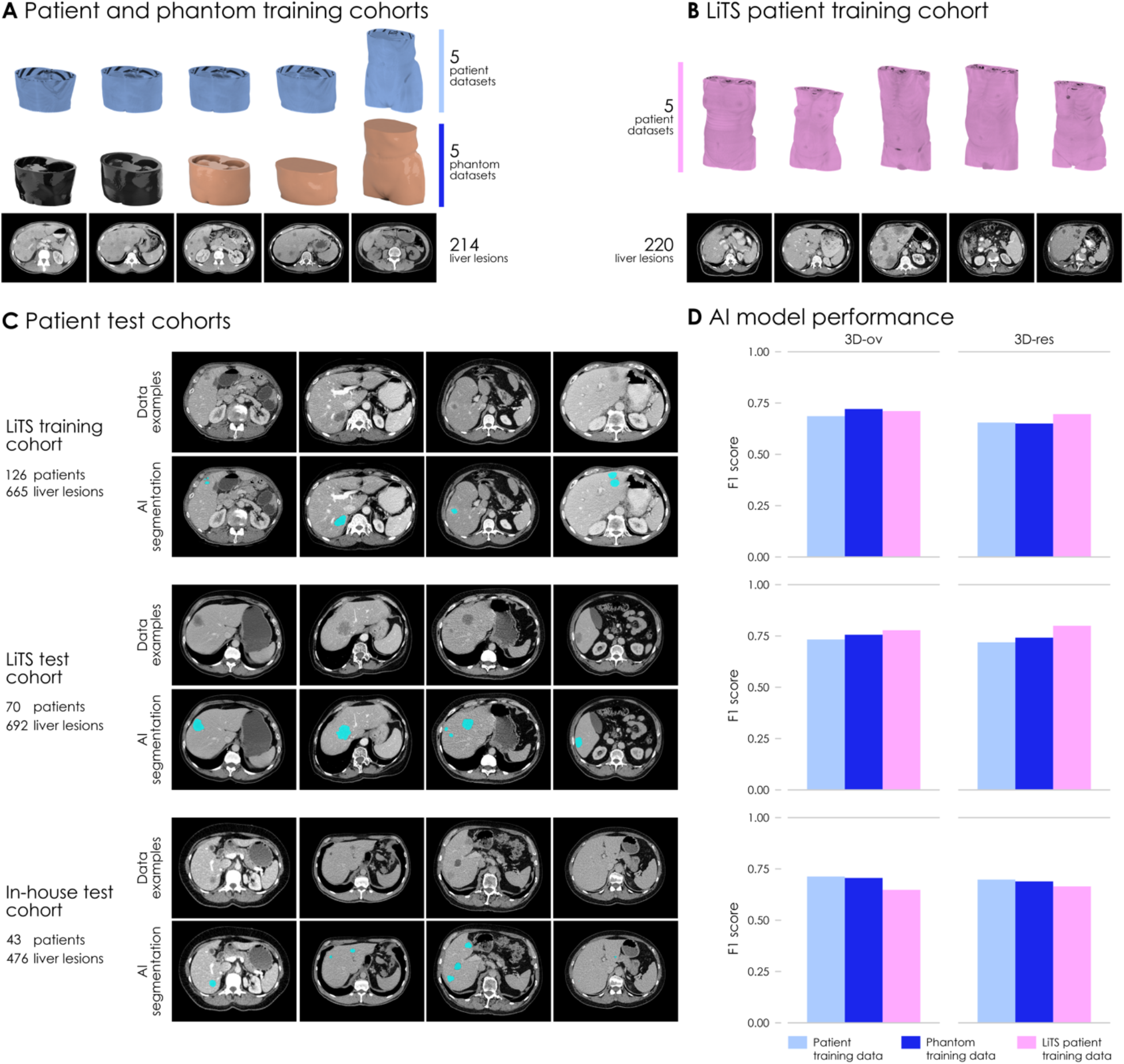
AI models trained on phantom-only and patient-only data. **a**, Patient and corresponding phantom training cohorts. Both cohorts consisted of the same five cases: two with naturally occurring liver lesions and three with synthetically inserted lesions. CT images shown originate from phantom scans. **b**, Subset of five cases from the Liver Tumor Segmentation (LiTS) training cohort with naturally occurring liver lesions. **c**, Clinical test cohorts used to evaluate AI models trained on the cohorts described in (a) and (b), including the remaining 126 cases from the LiTS training cohort, the LiTS test cohort, and an in-house test cohort. Images show example cases from each cohort with corresponding liver lesion segmentations generated by the original 3D full-resolution (3D-ov) U-Net trained on the phantom cohort. **d**, Liver lesion detection performance of U-Net models trained on patient-only, phantom-only and LiTS subcohort data, including both architectures: 3D-ov and a modified variant with residual connections in the encoder path (3D-res).

Lesion detection results showed that networks trained exclusively on phantom data achieved detection performance comparable to those trained on patient data when evaluated on clinical scans (**Fig. 5d**). Recall and precision were nearly identical between models trained on matching phantom and patient data (**Supplementary Fig. 8**). In contrast, models trained on the LiTS subcohort showed lower recall and higher precision. These trends were expected and reflect differences in lesion characteristics within the training data, the influence of which was amplified by the small number of training cases. Compared to the LiTS subcohort, models trained on phantom and associated patient data were exposed to lesions with lower conspicuity, resulting in greater sensitivity but also increased tendency to segment unlabeled areas in the test datasets. Numerical F1 score, recall and precision results are compiled in **Supplementary Table 5**.

Our findings demonstrate that AI models can effectively transfer patterns learned from phantom data to clinical application with performance comparable to models trained on real patient data. The results provide evidence that the phantoms matched the expected AI input domain and support their use in evaluating model performance. Moreover, they indicate the potential to also use the phantoms as a controlled source of prospective scan data to enrich AI training, which we explored in the final phase of our study.

### Enrichment of AI model training with phantom data

Training of diagnostic AI models typically relies on patient data, which consist of real scans but offer limited diversity in imaging conditions and little control over case characteristics. Synthetic data can be used to augment these datasets^27,28^, but they lack the physical realism of actual CT scans. Phantom-derived scans represent an alternative data source offering both physical realism and controlled acquisition. Building on our findings in the previous section, we used phantom-augmented training as a further test of clinical domain compatibility. To this end, we investigated whether supplementing patient training data with lesion-containing phantom scans could improve model performance, and confirmed through a lesion-free negative control that performance gains were driven by task-relevant features.

We supplemented the original 131 patient cases in the LiTS training dataset with 40 lesion-containing and 40 lesion-free phantom scans and trained both model architectures on each combined dataset (**Fig. 6a**). The lesion-containing scans originated from the two phantoms with naturally occurring liver lesions described in the previous section, and the lesion-free scans from two separate phantoms without liver lesions. The scans covered dose levels from 0.62 to 7.0 mGy and were reconstructed using HIR. Model performance was evaluated using the same test data as before: the 2,160 phantom scan datasets across both scanner systems and the clinical LiTS and in-house test cohorts. Results were then compared against the baseline of models trained on LiTS patient data alone.

**Fig. 6.**
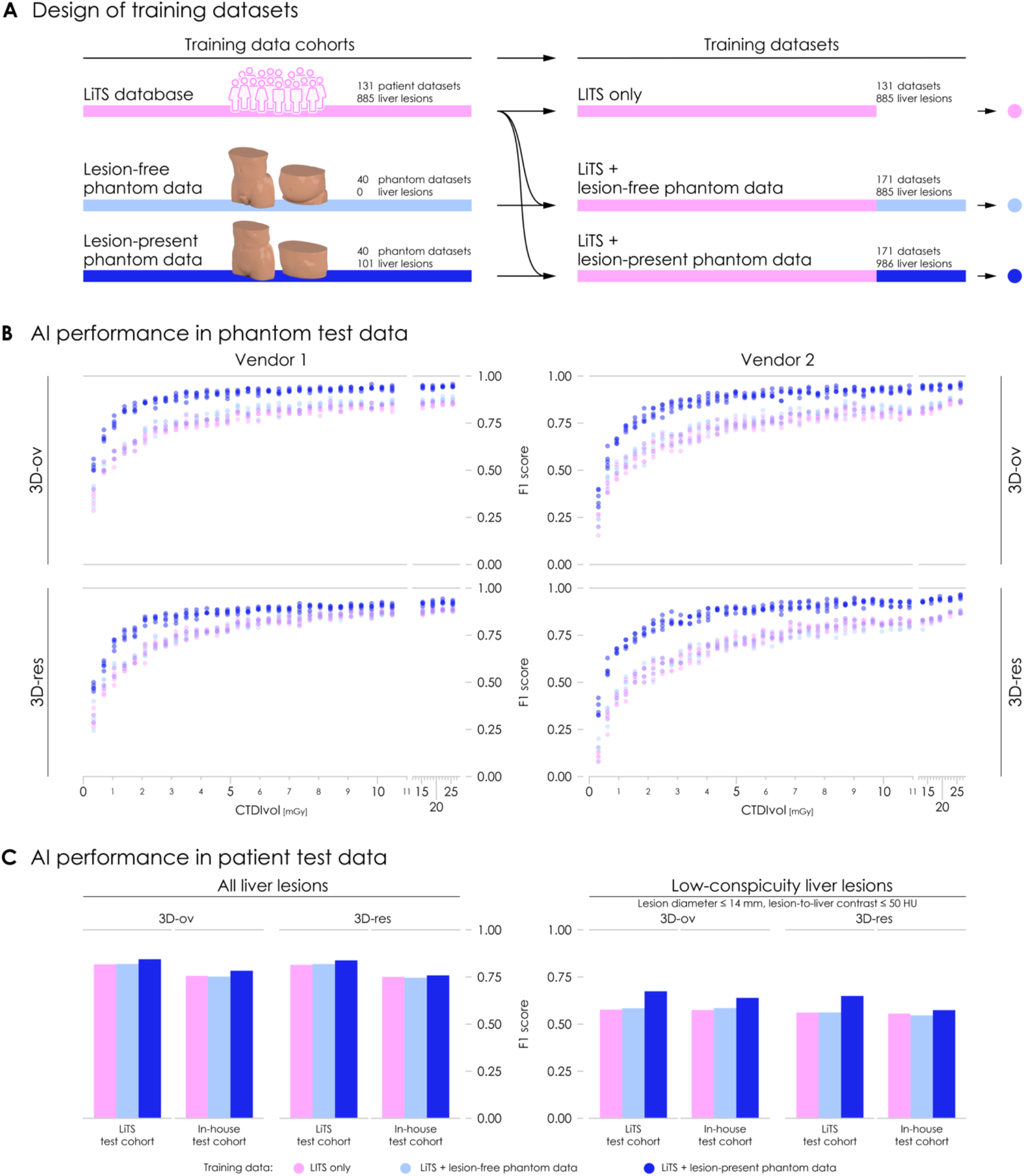
Phantom data-augmented AI training. **a**, Design of three training cohorts: the Liver Tumor Segmentation (LiTS) training cohort, the LiTS training cohort augmented with lesion-free phantom data, and the LiTS training cohort augmented with lesion-present phantom data from two phantoms containing a total of 101 liver lesions. Phantom data covered twenty scans per phantom across two CT scanners at dose levels ranging from 0.62 to 7.0 mGy. **b**, Liver lesion detection performance in phantom test data across both CT scanner systems and for U-Net models trained on the cohorts in (a), including the original 3D full-resolution (3D-ov) and a modified variant with residual connections in the encoder path (3D-res). CTDIvol is the volume computed tomography dose index. **c**, Liver lesion detection performance in clinical LiTS and in-house test cohorts across all lesions, with a subanalysis for low-conspicuity lesions 14 mm or smaller in diameter and -50 HU or greater lesion-to-liver contrast.

**Figure 6b** shows detection results on the phantom test data. Models trained on the combined lesion-containing dataset consistently outperformed those trained on patient data alone across both scanners and all dose levels. The largest absolute and relative gains were observed at lower doses, while improvements were more modest at high doses where baseline performance was already strong. The improvement in F1 scores was driven by a strong increase in recall, with a marginal decrease in precision (**Supplementary Fig. 9**).

The lesion-free control experiment confirmed that these improvements were not due to domain adaptation. When phantom scans without lesions were added to the training data, no performance change was observed compared to models trained on patient data only. This finding, together with our earlier experiments, confirms that any phantom-specific imaging characteristics were either absent or irrelevant to model learning. Instead, performance gains were driven by task-relevant features introduced by the phantom scans.

Improved model performance through phantom data-augmented training was confirmed in the clinical test cohorts, with the most pronounced gains observed for low-conspicuity lesions whose characteristics matched those of the phantom training data (**Fig. 6c**). Subanalysis by lesion type in the phantom test data confirmed this pattern. The strongest improvements occurred for the least conspicuous lesions, whereas the most conspicuous lesions, which already exhibited near-perfect baseline detection, showed little change (**Fig. 7**). Consistent with the recall-precision trade-off described earlier, improvements in F1 scores were driven by an increase in recall, with precision decreasing slightly or remaining near constant (**Supplementary Fig. 10 and 11**).

**Fig. 7.**
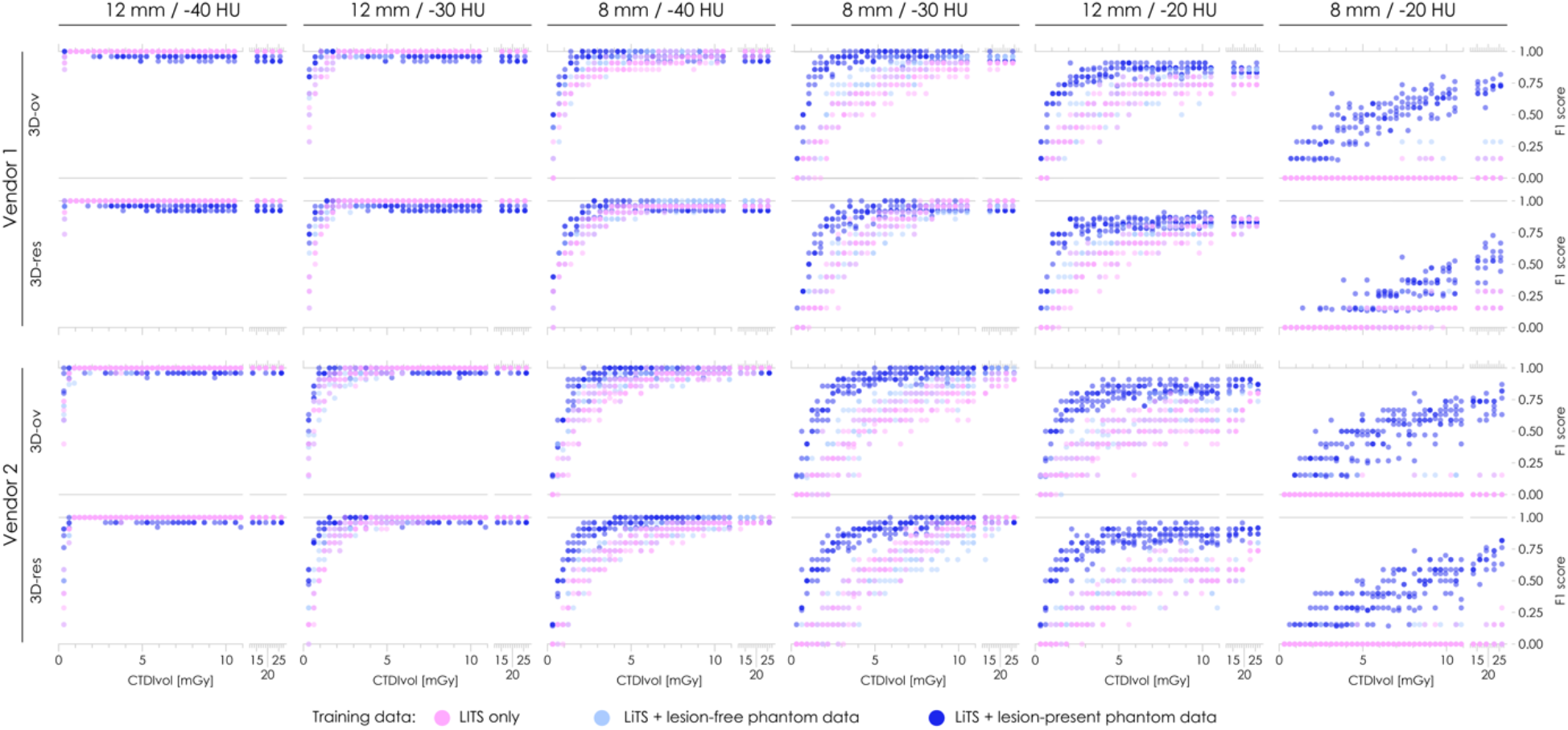
AI detection performance across liver lesion subgroups after phantom-augmented training. Results are presented for for each lesion diameter and lesion-to-liver contrast combination labelled at the top of each panel across both CT scanner systems and nnU-Net model architectures: the original 3D full-resolution configuration (3D-ov) and a modified variant with residual connections in the encoder path (3D-res). CTDIvol is the volume computed tomography dose index.

Our results demonstrate that phantom scans can effectively supplement patient training data to improve AI model performance. Gains were driven by task-relevant lesion features rather than phantom-specific imaging characteristics, further confirming clinical domain compatibility. As real scan data that can be prospectively collected and controlled in terms of case characteristics and imaging conditions, phantoms can additionally offer a targeted means to address specific performance gaps in AI model development. Our findings represent an initial demonstration of this potential, with a more systematic exploration of training strategies warranting further investigation.

## Discussion

As AI becomes more deeply embedded in imaging workflows, robust mechanisms to ensure consistent, safe, and trustworthy deployment are becoming increasingly important. Efforts are underway to formalize AI oversight, including accreditation programs led by professional bodies such as the American College of Radiology^29^. These initiatives aim to establish systematic frameworks for ongoing evaluation and quality assurance. However, methods for local, on-site validation and surveillance of scan data quality and AI functionality remain largely limited to retrospective evaluation. Current monitoring strategies rely heavily on performance feedback during clinical use and are inherently constrained by their reactive nature and the ethical and practical limitations of testing in patients.

Here, we present a framework for standardised on-site testing and monitoring of AI performance. We design physical phantoms that match the expected anatomical input domain of AI models and serve as consistent imaging targets for reproducible testing. Using these tools, we systematically assess AI performance across two clinical CT scanners, demonstrating sensitivity to variations in dose, image reconstruction, scanner technology, model architecture, and lesion characteristics. We establish a monitoring strategy and demonstrate its application for longitudinal evaluation. To validate clinical relevance, we show that AI models trained on phantom data exhibit no signs of phantom-specific adaptation and transfer effectively to patient cases. Furthermore, we demonstrate that phantom-derived scan data can enrich AI training for improved performance.

Phantom-based, end-to-end validation of imaging and AI workflows extends conventional phantom quality assurance beyond basic scanner-focused assessment. By incorporating realistic anatomical and task-specific features, it enables systematic evaluation of AI functionality in a controlled and reproducible setting. Prior work has described related concepts, including the use of realistic phantoms^30^ and clinically predictive testing^31^. Building on these advances, our framework integrates anatomically realistic phantoms with AI assessment to create a standardised and proactive mechanism for testing, longitudinal monitoring, and revalidation.

Applying this framework, we systematically characterized the effects of key imaging parameters on the performance of two AI models. We observed pronounced sensitivity to dose variations, particularly within the range of typical oncologic CT protocols^32^, as well as improvements associated with denoising image reconstruction and differences across scanner technologies. An update to the AI model architecture was found to interact with scanner type and dose in influencing performance. By providing a consistent reference, the framework makes these sources of variability transparent and quantifiable. Longitudinal measurements further confirmed that AI behaviour remained stable under unchanged conditions, demonstrating that the framework supports both the detection of performance variability and ongoing monitoring of system consistency over time.

Such testing cannot be performed in patients. To validate clinical relevance, we therefore demonstrate that the AI models interpret phantom images as valid imaging input and effectively transfer patterns learned from phantom data to patient cohorts. In these validation experiments, improved model performance indicates additional potential of the framework as a controlled source for generating AI training data. Because such data can be prospectively acquired and include precise ground truth, this approach enables targeted augmentation of training datasets and controlled exposure of AI models to imaging characteristics. While our observations provide proof of principle for this application, a more comprehensive evaluation of optimal training strategies warrants further investigation.

Liver lesion detection served as a demonstration task here, but the underlying framework is not limited to a single organ, task, or imaging modality, as phantom designs are inherently adaptable to different target applications^14,33^. Reported applications of task-matched phantom designs for AI-based hepatic lesion classification^34^, intracranial aneurysm detection^35^, and breast cancer screening^36^ in CT and mammography reflect this adaptability. Corroborated by these initial applications, our findings provide evidence supporting the framework as a broadly applicable quality assurance method for site-specific evaluation and monitoring of AI image analysis models.

Finally, it should be noted that the framework presented here is intended to evaluate AI functionality as a function of scan data quality rather than providing an absolute measure of AI performance across biological variability. Phantoms provide references for assessing imaging and AI performance, but they are not primarily intended to replace large patient cohorts with diverse anatomies and pathologies. Patient datasets therefore remain essential for validating AI efficacy across populations, while a phantom-based framework provides a technical solution for on-site validation, longitudinal monitoring, and cross-institutional comparison. In this context, it also provides a pathway toward improved harmonization of diagnostic imaging quality across institutions, where substantial variation still persists^37^.

In conclusion, we present a phantom-based framework for standardised, prospective testing and monitoring of AI image analysis across scanner systems. By using anatomically and task-matched reference phantoms, the framework makes performance variability visible, measurable, and traceable at the local level. It enables consistent quality assurance and can support harmonization of imaging standards across institutions. Beyond evaluation, it also offers a controlled source of imaging data for targeted AI training, with its broader impact on model development yet to be established.

## Methods

### Phantoms

Institutional ethics committee approval was obtained for using CT images of patients to construct anatomically realistic phantoms and for analysing diagnostic performance in patient and phantom image data. Lesion-free abdominal CT data covering the entire liver were extracted from two portal-venous phase CT datasets^38^. Spherical lesions with diameters of 8 or 12 mm and nominal contrasts of -10, -20, -30, or -40 HU relative to the surrounding liver tissue were digitally inserted. Six lesions were inserted for each size-contrast combination, except for -10 HU contrast lesions, which were only inserted with a diameter of 12 mm. Lesions were distributed across seven sections in Phantom 1 and six sections in Phantom 2. The resulting data were used to produce two physical phantoms using a technology based on previously established methods^17,18^, in which CT image data are first inkjet-printed layer by layer using iodine-doped ink onto cellulose sheets, followed by three-dimensional assembly of the printed layers. Both phantoms were manufactured by PhantomX GmbH (Berlin, Germany). Phantom 1 (manufacturer model name PX0203542) measured 268 x 189 x 150 mm, and Phantom 2 (PX0203444c) measured 247 x 186 x 135 mm. Three additional phantoms used for comparative training and evaluation of AI models trained on phantom-only and patient-only data were phantoms PX0203547, PX0203548, and PX0203445 supplied by PhantomX. Phantoms PX0203548 and PX0203445 were subsequently also employed in phantom-augmented training to enrich the LiTS patient training dataset. Two additional phantoms (PX0203544 and PX0203444) were used for the negative control.

### AI network training

The nnU-Net framework was used to train two 3D U-Net models for semantic segmentation of the liver and liver lesions^23,24^. The framework provides standardised guidelines and predefined configurations, from which the best-performing non-ensemble setup was selected. This included the 3D full resolution setup in its original version (3D-ov) and the most recent ResEnc L version incorporating residual encoders (3D-res). Prior to training all data were converted to the NIfTI format using the Python package NiBabel (v5.1.0). Data normalization followed the recommended nnU-Net procedure for CT images. In brief, voxel intensities corresponding to liver and liver lesion regions were collected across all training cases to compute the mean, standard deviation, and the 0.5 and 99.5 percentiles. Intensity values were clipped to this percentile range and subsequently normalized by subtracting the mean and dividing by the standard deviation. In accordance with nnU-Net guidelines the networks were trained using five fold cross validation. Each training run lasted 1000 epochs. The models from the final training epoch were used for subsequent network evaluation.

### AI network training data

AI models were trained on the full training dataset released within the Liver Tumor Segmentation (LiTS) challenge, consisting of 131 abdominal patient scans and the corresponding segmentation masks. The dataset contained 885 liver lesions and 58638 images in total, was collected from seven clinical sites across five countries, and is publicly available in NIfTI format^39^.

### Patient-only and phantom-only training data

For the phantom-only dataset, images acquired from five phantoms were used: Phantom 1 (PX0203542), Phantom 2 (PX0203444c), and phantoms PX0203547, PX0203548, and PX0203445. Scans were acquired on a Canon Aquilion One CT scanner (Canon Medical Systems, Otawara, Japan) at 5.25 mGy and reconstructed with adaptive iterative dose reduction 3D (AIDR 3D, FC08 kernel), with a voxel spacing of 0.59 x 0.59 x 0.8 mm. The corresponding patient-only dataset consisted of the patient data used to generate these five phantoms. The second patient-only training set was a subset of five patients from the LiTS training dataset (volumes 49, 93, 108, 109, and 117). This subset comprised 3398 images with voxel spacing ranging from 0.66 x 0.66 x 0.8 mm to 0.96 x 0.96 x 2.5 mm.

### Phantom data-augmented training data

The phantom data-augmented training dataset consisted of the full LiTS training dataset and additional lesion-present phantom scans from phantoms PX0203548 and PX0203445 and lesion-free phantom scans from phantoms PX0203544 and PX0203444 (PhantomX GmbH). The lesion-present phantoms PX0203548 and PX0203445 contained a total of 101 liver lesions. Phantom scans were acquired in equal numbers on two CT scanner systems: a Canon Aquilion One using AIDR 3D reconstruction with the FC08 kernel and a voxel spacing of 0.59 x 0.59 x 1 mm, and a GE Revolution using Adaptive Statistical Iterative Reconstruction (ASIR) with the Standard kernel and a voxel spacing of 0.59 x 0.59 x 0.625 mm. Ten scans were performed per phantom and scanner system across dose levels of 0.7 to 7.0 mGy on the Canon scanner and 0.62 to 6.2 mGy on the GE scanner, in increments of 0.7 mGy and 0.62 mGy respectively.

### Ground truth masks in phantom scans

Phantom lesion segmentation ground-truth masks in scans of Phantom 1 (PX0203542) and Phantom 2 (PX0203444c) were generated programmatically using Python (v3.9.11) based on the lesion location information and diameters specified during phantom design. In a first step, lesion coordinates and diameters were used to generate ground-truth masks for the image data used as template for the physical phantoms using the Python packages NumPy v1.26.4 and SimpleITK v2.2.1. These masks were reviewed for accuracy and subsequently transferred to the phantom scans via image registration using the Insight Segmentation and Registration Toolkit (ITK) and its Python wrapper (v5.3.0, Python v3.9.11), as well as its extension itk-elastix (v0.17.1). For phantoms PX0203547, PX0203548, and PX0203445, lesions were segmented using ITK-SNAP (v4.2.2) by a radiologist in training with 3 years of experience and subsequently reviewed by a board-certified radiologist with 12 years of experience using scans acquired at 25.5 mGy on a Canon Aquilion One CT scanner reconstructed with AIDR 3D (FC08 kernel).

### Phantom test data

For AI model evaluation and monitoring, Phantom 1 and Phantom 2 were scanned on a Canon Aquilion One (Canon Medical Systems, Otawara, Japan) referred to as Vendor 1 and a GE Revolution (GE Healthcare, Milwaukee, USA) referred to as Vendor 2. A tube voltage of 120 kVp was employed on both scanners, and the tube current and rotation time were varied to adjust the volume computed tomography dose index (CTDIvol) between 0.35 and 25.50 mGy for Vendor 1 and between 0.31 and 26.1 mGy for Vendor 2. All available image reconstruction algorithms were used including filtered back projection (FBP) and hybrid iterative reconstruction (HIR) on both scanners and additionally model-based iterative reconstruction (MBIR) and deep learning reconstruction (DLR) on Vendor 1. The vendor-specific implementations were AIDR 3D (Vendor 1) and ASIR (Vendor 2) for HIR, Forward projected model-based Iterative Reconstruction SoluTion (FIRST) for MBIR and Advanced intelligent Clear-IQ Engine (AiCE) for DLR. Five repeated acquisitions were performed per dose and reconstruction method. The scanners’ standard abdominal image slice thickness was applied (1 mm for Vendor 1 and 0.625 mm for Vendor 2). **Supplementary Table 1** provides a detailed overview of scan settings. To adjust the slice thickness of Vendor 2 images for the comparative analysis between scanner systems, images were linearly interpolated to 1 mm using Python version 3.9.11 and the Python frameworks numpy (v1.26.4) and pydicom (v2.4.1).

### Patient test data

For the comparative evaluation of patient-only and phantom-only trained AI models, three patient test datasets were employed. The first dataset consisted of 126 cases from the LiTS training dataset excluding the five cases used for model training. This dataset comprised 665 liver lesions with a median volume of 681 mm^3^ (range 1.2 mm^3^ to 968636.2 mm^3^) and a median lesion to liver contrast of -37 HU (range -141 HU to 42 HU). The second dataset was the test dataset released as part of the LiTS challenge and consisted of 70 cases with 692 liver lesions. The median lesion volume was 784 mm^3^ (range 1.5 mm^3^ to 1233596 mm^3^) and the median lesion to liver contrast was -31 HU (range -100 HU to 87 HU). The third dataset consisted of 43 cases collected in-house across CT scanners from three manufacturers detailed in **Supplementary Table 4** and comprised 476 liver lesions. The median lesion volume was 405 mm^3^ (range 1.2 mm^3^ to 457491 mm^3^) and the median lesion to liver contrast was -28 HU (range -90 HU to 66 HU). Ground truth liver lesion segmentation masks for the LiTS test dataset and the in-house test dataset were created by a radiologist in training with 3 years of experience and reviewed by a board certified radiologist with 12 years of experience.

### Lesion subgroup analysis

For the calculation of detection metrics on lesion subgroups, the contrast and volumes of all ground truth and predicted lesion masks were determined. Lesion volume was calculated by multiplying the number of voxels within each lesion by the voxel volume, which was derived from the pixel spacing. To evaluate lesion contrast relative to the surrounding liver tissue, the segmentation masks were dilated by nine pixels in all directions using Python scripts based on the libraries SciPy (v1.13.1) and NiBabel (v5.1.0). Lesion contrast was calculated by subtracting the mean HU value of the lesion mask from the mean HU value of the seven outermost layers of the dilated mask using the Python libraries numpy (v1.26.4) and SimpleITK (v2.2.1). To avoid inclusion of voxels belonging to other lesions or voxels outside the liver tissue, the dilated masks were cleaned using liver segmentation masks and ground truth lesion segmentation masks.

### Assessment of AI lesion detection performance

Lesion detection was assessed by comparing AI-generated segmentation masks with ground-truth lesion masks. Overlapping segmentations were classified as true positives (TP), whereas AI segmentations without overlap with the ground truth were considered false positives (FP) and ground-truth segmentations without overlap with AI predictions were considered false negatives (FN). Lesions were defined as three-dimensional connected components of the segmentation masks. To establish correspondence between predicted and reference lesions while accounting for potential under-segmentation (one prediction covering multiple lesions) and over-segmentation (multiple predictions corresponding to a single lesion), a previously described matching procedure was applied^25^. Briefly, ground truth lesions connected through a single predicted component were merged into a single component. If a ground truth lesion overlapped multiple predicted components, only the predicted component with the largest overlap was retained. Ground truth components without any overlapping prediction were classified as FN. In a second step, each remaining predicted component was associated with the ground truth component from the first step with which it had the largest intersection. Predicted components associated with the same ground truth component were merged. Predicted components without any corresponding ground truth component were classified as FP. This procedure produced correspondence maps between sets of predicted and ground truth components. Metrics calculated for merged ground truth components were subsequently attributed to the individual ground truth lesions. A lesion was considered a TP if the predicted mask overlapped with the corresponding ground truth lesion, defined as a Dice coefficient greater than 0. Precision and recall were calculated as TP/(TP+FP) and TP/(TP+FN), respectively, and their harmonic mean was reported as the F1 score. TP, FP, and FN were summed across all cases before computing precision, recall, and F1 to account for cases without lesions. Masks with a volume smaller than that of a sphere with 5 mm diameter (65.45 mm^3^) were excluded from the analysis. For the subgroup analysis, masks were included if they had a contrast between -50 HU and 0 HU and a volume between 65.45 mm³ and 1436.76 mm³ (equivalent to spheres with diameters of 5 mm and 14 mm, respectively).

### Statistical analysis

Group comparisons of AI liver lesion detection performance across reconstruction algorithms, scanner systems, and network architectures were performed descriptively by comparing the area under the curve (AUC) of F1 score-dose curves. The AUC was calculated over the dose range of 0.35-10.5 mGy for Vendor 1 and 0.31-10.88 mGy for Vendor 2. Higher dose levels were excluded due to their limited clinical relevance, lower sampling density, and the plateauing of performance curves at these levels. AUC values were normalized by dividing the raw AUC by the theoretical maximum area, corresponding to perfect detection performance across all dose levels.

## Data Availability

The main data supporting the results in this study are available within the paper and its Supplementary Information.

## Acknowledgements

Computation has been performed on the HPC for Research cluster of the Berlin Institute of Health.

## Supplementary Information

### 1. AI liver lesion detection performance

#### 1.1 Phantom test data acquisition parameters

**Supplementary Table 1.** CT acquisition parameters for phantom test data collection. Scan settings and resulting volume computed tomography dose indices (CTDIvol) used to scan Phantom 1 and Phantom 2 across both scanner systems for AI model performance evaluation.

| CT system | Tube voltage (kVp) | Rotation time (s) | Tube current (mA) | Pitch | CTDIvol (mGy) |
| --- | --- | --- | --- | --- | --- |
| Vendor 1 | 120 | 0.5 | 10, 20, 30, 40, 50, 60, 70, 80, 90, 100, 110, 120, 130, 140, 150, 160, 170, 180, 190, 200, 210, 220, 230, 240, 250, 260, 270, 280, 290, 300, 400, 500, 600, 700 | 0.813 | 0.35, 0.7, 1.05, 1.4, 1.75, 2.1, 2.45, 2.8, 3.15, 3.5, 3.85, 4.2, 4.55, 4.9, 5.25, 5.6, 5.95, 6.3, 6.65, 7, 7.35, 7.7, 8.05, 8.4, 8.75, 9.1, 9.45, 9.8, 10.15, 10.5, 14.6, 18.23, 21.86, 25.49 |
| Vendor 2 | 120 | 0.5 | 10, 20, 30, 40, 50, 60, 70, 80, 90, 100, 110, 120, 130, 140, 150, 160, 170, 180, 190, 200, 210, 220, 230, 240, 250, 260, 270, 280, 290, 300, 310, 320, 330, 340, 350, 400, 500, 600 | 0.99 | 0.31, 0.62, 0.93, 1.24, 1.55, 1.87, 2.18, 2.49, 2.80, 3.11, 3.42, 3.73, 4.04, 4.35, 4.66, 4.98, 5.29, 5.60, 5.91, 6.22, 6.53, 6.84, 7.15, 7.46, 7.77, 8.08, 8.40, 8.71, 9.02, 9.33, 9.64, 9.95, 10.26, 10.57, 10.88, 12.44, 15.55, 18.66, 22.39, 26.12 |
| Vendor 2 | 120 | 0.6 | 600 | 0.99 | 22.39 |
| Vendor 2 | 120 | 0.7 | 600 | 0.99 | 26.12 |

#### 1.2 Recall and precision

**Supplementary Fig. 1.**
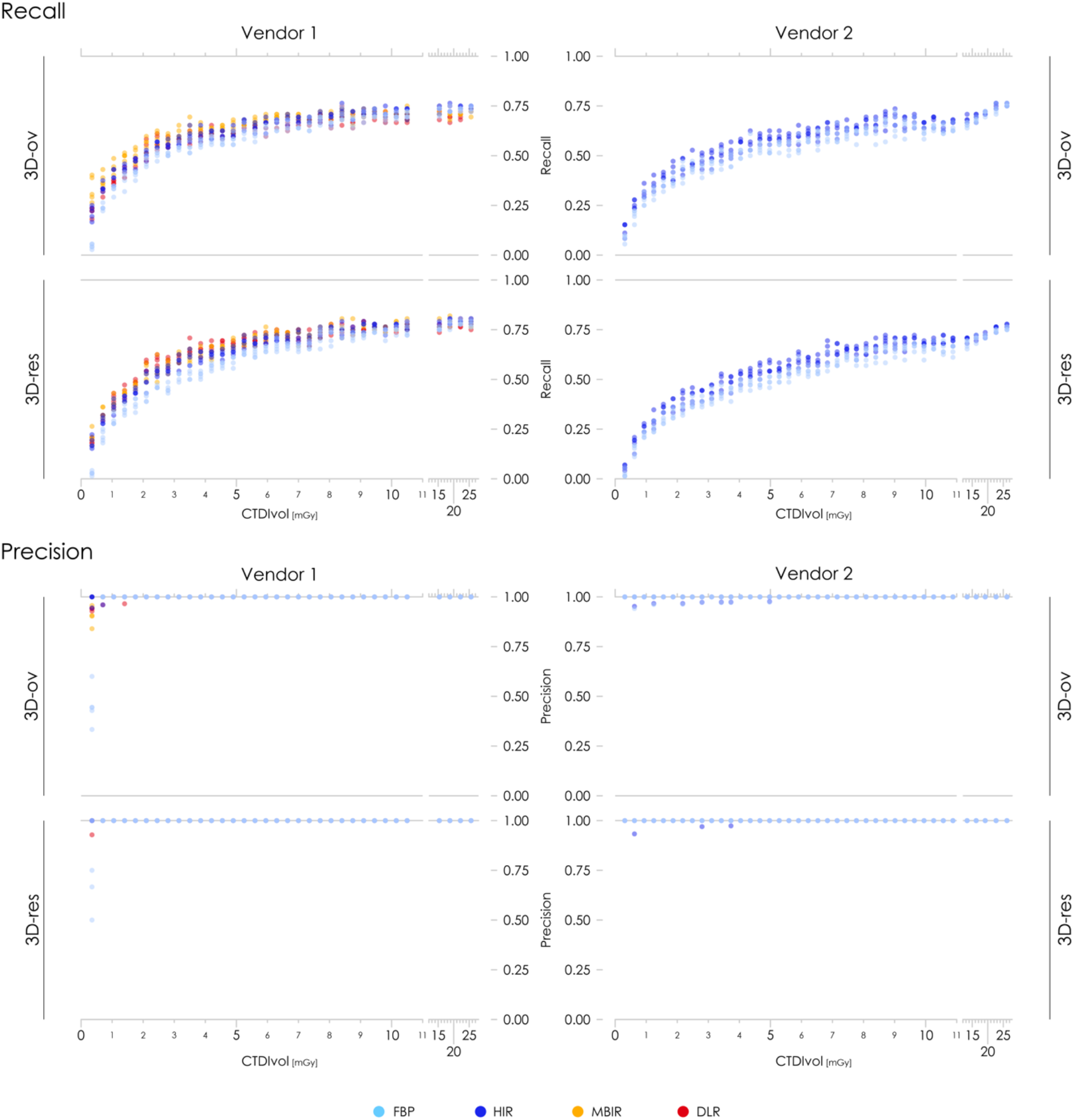
Recall and precision of AI-based liver lesion detection. Results are shown for both nnU-Net architectures, the original 3D full-resolution configuration (3D-ov) and the modified variant with residual encoder connections (3D-res), both scanner systems, all dose levels, five repeated acquisitions, and all reconstruction algorithms: filtered back projection (FBP), hybrid iterative reconstruction (HIR), model-based iterative reconstruction (MBIR), and deep learning reconstruction (DLR). CTDIvol is the volume computed tomography dose index.

### 2. Comparison of scanner technologies and AI architectures

#### 2.1 Numerical results

**Supplementary Table 2.** Area under the curve (AUC) values of F1 scores. Mean AUC and ranges across dose levels below 11 mGy and five repeated acquisitions are presented for both scanner systems, both nnU-Net architectures, the original 3D full-resolution configuration (3D-ov) and the modified variant with residual encoder connections (3D-res), and all reconstruction algorithms: filtered back projection (FBP), hybrid iterative reconstruction (HIR), model-based iterative reconstruction (MBIR), and deep learning reconstruction (DLR). Resliced Vendor 2 results refer to F1 scores computed on Vendor 2 data linearly interpolated to match the slice thickness of Vendor 1.

| CT system | nnU-Net architecture | Image reconstruction algorithm | AUC (mean) | AUC (range) |
| --- | --- | --- | --- | --- |
| Vendor 1 | 3D-ov | DLR | 0.718 | 0.716 - 0.722 |
| Vendor 1 | 3D-ov | FBP | 0.691 | 0.686 - 0.692 |
| Vendor 1 | 3D-ov | HIR | 0.726 | 0.720 - 0.729 |
| Vendor 1 | 3D-ov | MBIR | 0.743 | 0.739 - 0.746 |
| Vendor 1 | 3D-res | DLR | 0.751 | 0.748 - 0.755 |
| Vendor 1 | 3D-res | FBP | 0.689 | 0.688 - 0.692 |
| Vendor 1 | 3D-res | HIR | 0.734 | 0.731 - 0.737 |
| Vendor 1 | 3D-res | MBIR | 0.747 | 0.744 - 0.753 |
| Vendor 2 | 3D-ov | FBP | 0.653 | 0.649 - 0.658 |
| Vendor 2 | 3D-ov | HIR | 0.685 | 0.683 - 0.688 |
| Vendor 2 | 3D-res | FBP | 0.626 | 0.620 - 0.630 |
| Vendor 2 | 3D-res | HIR | 0.665 | 0.659 - 0.670 |
| Vendor 2 resliced | 3D-ov | HIR | 0.676 | 0.672 - 0.681 |
| Vendor 2 resliced | 3D-res | HIR | 0.633 | 0.629 - 0.639 |

#### 2.2 Image reconstruction algorithms

**Supplementary Fig. 2.**
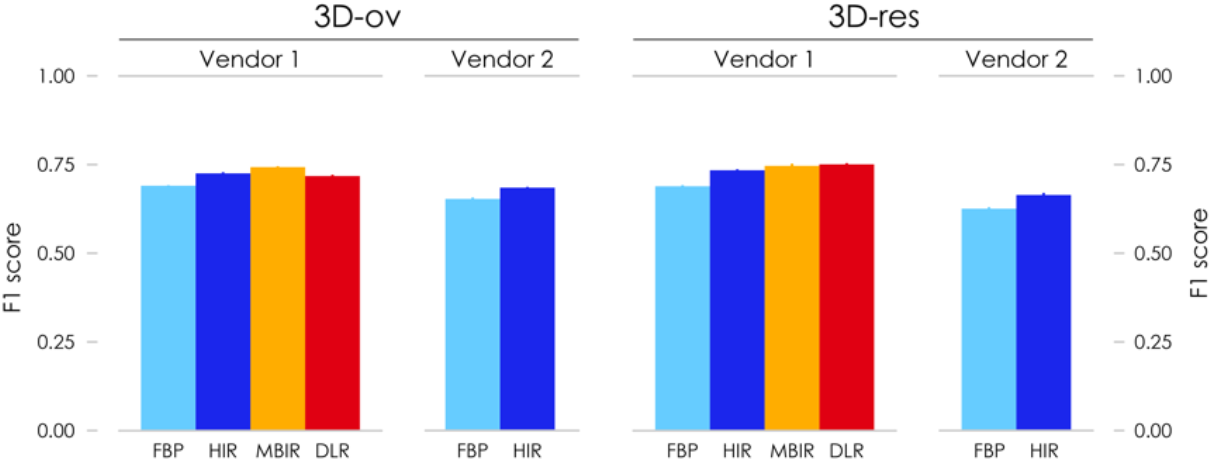
Area under the curve of F1 scores across image reconstruction algorithms. Mean results across dose levels below 11 mGy and five repeated acquisitions are shown for both nnU-Net architectures, the original 3D full-resolution configuration (3D-ov) and the modified variant with residual encoder connections (3D-res), both scanner systems, and all reconstruction algorithms: filtered back projection (FBP), hybrid iterative reconstruction (HIR), model-based iterative reconstruction (MBIR), and deep learning reconstruction (DLR). Error bars indicate ranges across five repeated acquisitions.

#### 2.3 Scanner models

**Supplementary Fig. 3.**
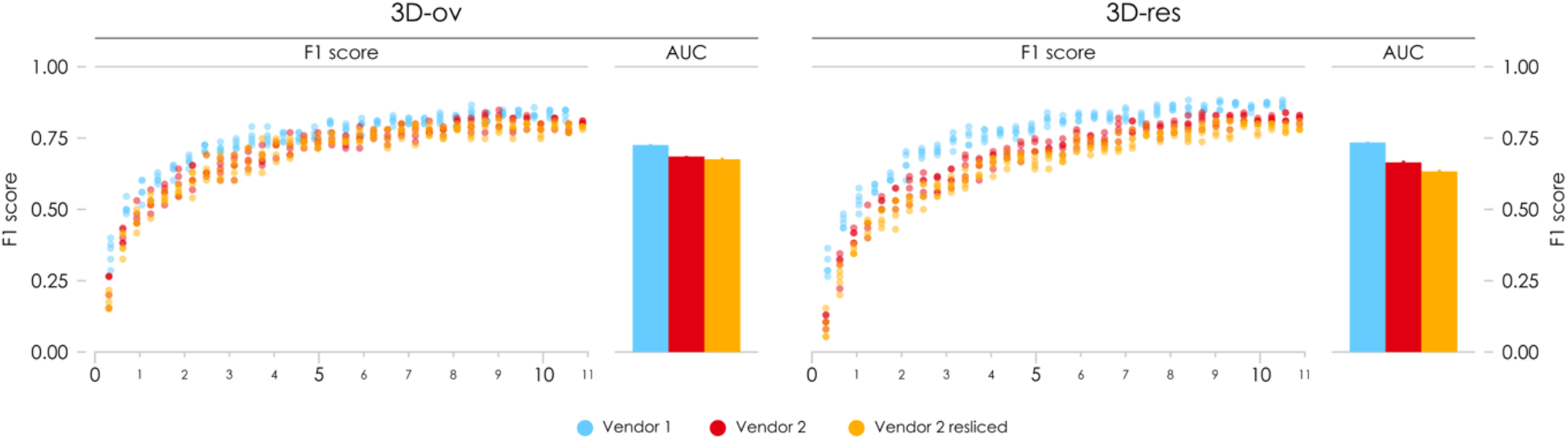
F1 scores and area under the curve (AUC) across scanner systems. F1 scores and mean AUC across dose levels below 11 mGy and five repeated acquisitions are shown for both nnU-Net architectures, the original 3D full-resolution configuration (3D-ov) and the modified variant with residual encoder connections (3D-res), and both scanner systems. Resliced Vendor 2 results refer to F1 scores and AUC values computed on Vendor 2 data linearly interpolated to match the slice thickness of Vendor 1. CTDIvol is the volume computed tomography dose index. Error bars indicate ranges across five repeated acquisitions.

#### 2.4 AI architectures

**Supplementary Fig. 4.**
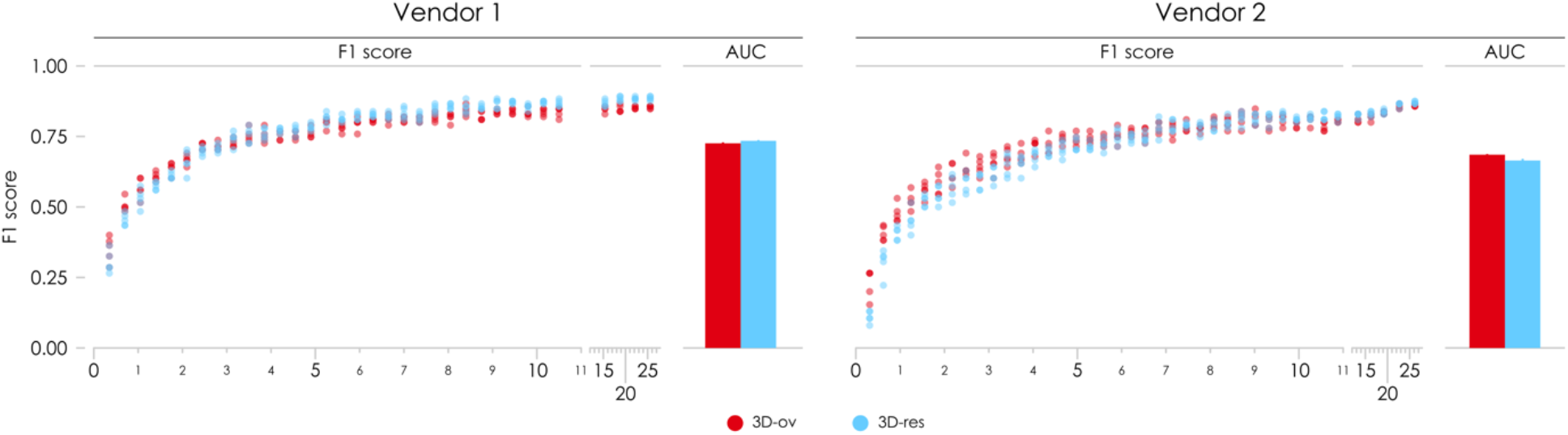
F1 scores and area under the curve (AUC) across AI model architectures. F1 scores and mean AUC across dose levels below 11 mGy and five repeated acquisitions are shown for both nnU-Net architectures, the original 3D full-resolution configuration (3D-ov) and the modified variant with residual encoder connections (3D-res), and both scanner systems. CTDIvol is the volume computed tomography dose index. Error bars indicate ranges across five repeated acquisitions.

### 3. Lesion subanalysis

#### 3.1 Recall and precision

**Supplementary Fig. 5.**
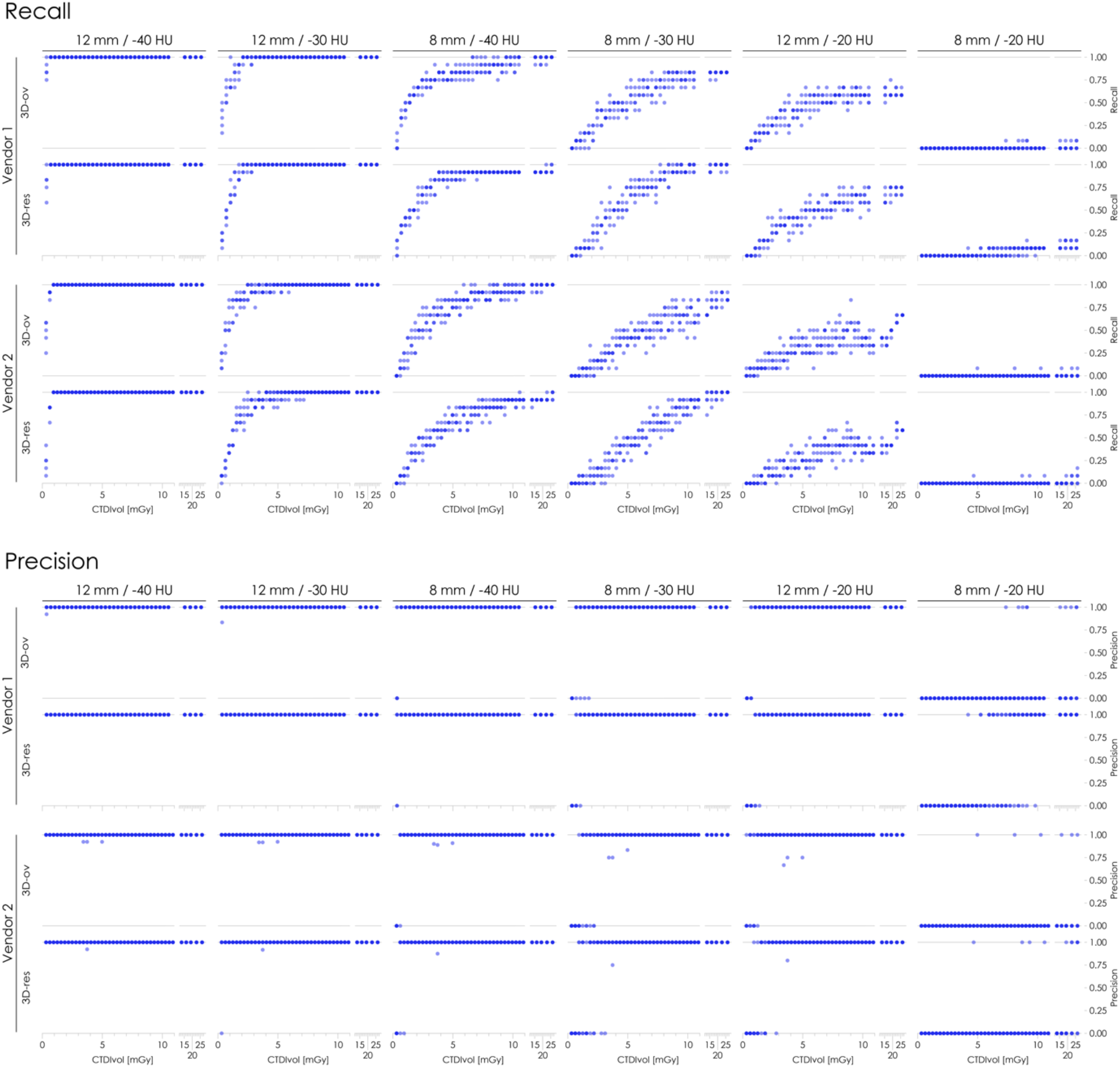
Recall and precision of AI-based liver lesion detection across liver lesion subgroups. Results are presented for for each lesion diameter and lesion-to-liver contrast combination labelled at the top of each panel across both scanner systems and nnU-Net architectures: the original 3D full-resolution configuration (3D-ov) and the modified variant with residual encoder connections (3D-res). CTDIvol is the volume computed tomography dose index.

### 4. Longitudinal AI monitoring

#### 4.1 Recall and precision

**Supplementary Fig. 6.**
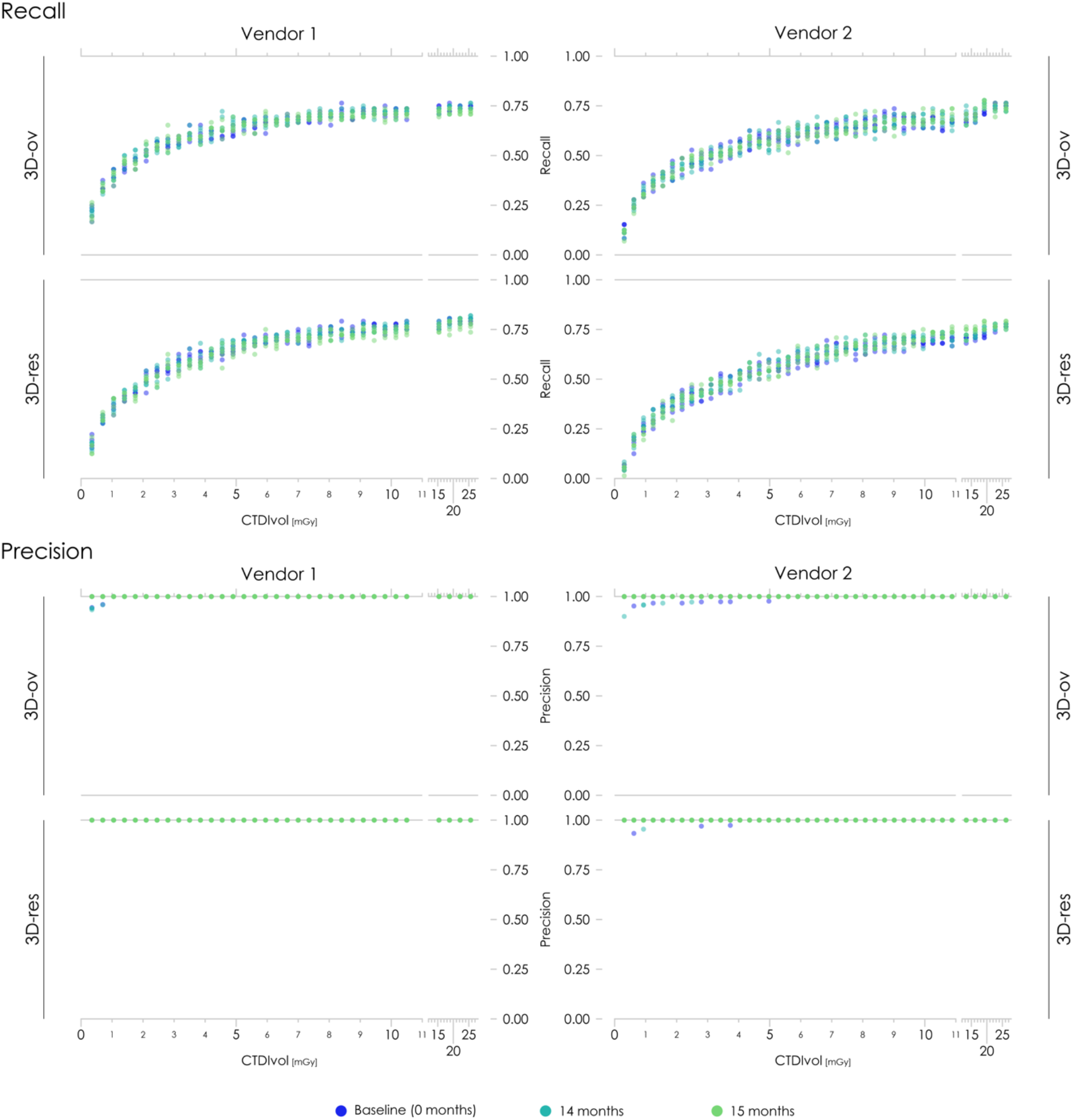
Recall and precision during longitudinal monitoring. Results are shown for both nnU-Net architectures, the original 3D full-resolution configuration (3D-ov) and the modified variant with residual encoder connections (3D-res), both scanner systems, all dose levels, and five repeated acquisitions. CTDIvol is the volume computed tomography dose index.

#### 4.2 Numerical results

**Supplementary Table 3.** Area under the curve (AUC) values of F1 scores across longitudinal monitoring time points. Mean AUC and ranges across volume computed tomography dose indices (CTDIvol) below 11 mGy and five repeated acquisitions are presented for both scanner systems and both nnU-Net architectures, the original 3D full-resolution configuration (3D-ov) and the modified variant with residual encoder connections (3D-res), at baseline (0 months) and after 14 and 15 months.

| CT system | nnU-Net architecture | CTDIvol range (mGy) | Time point (months) | AUC (mean) | AUC (range) |
| --- | --- | --- | --- | --- | --- |
| Vendor 1 | 3D-ov | 0.35 - 10.5 | 0 | 0.726 | 0.720 - 0.729 |
| Vendor 1 | 3D-ov | 0.35 - 10.5 | 14 | 0.732 | 0.730 - 0.734 |
| Vendor 1 | 3D-ov | 0.35 - 10.5 | 15 | 0.727 | 0.725 - 0.730 |
| Vendor 1 | 3D-res | 0.35 - 10.5 | 0 | 0.734 | 0.731 - 0.737 |
| Vendor 1 | 3D-res | 0.35 - 10.5 | 14 | 0.736 | 0.732 - 0.737 |
| Vendor 1 | 3D-res | 0.35 - 10.5 | 15 | 0.725 | 0.721 - 0.728 |
| Vendor 2 | 3D-ov | 0.31 - 10.88 | 0 | 0.685 | 0.683 - 0.688 |
| Vendor 2 | 3D-ov | 0.31 - 10.88 | 14 | 0.687 | 0.682 - 0.694 |
| Vendor 2 | 3D-ov | 0.31 - 10.88 | 15 | 0.684 | 0.682 - 0.688 |
| Vendor 2 | 3D-res | 0.31 - 10.88 | 0 | 0.665 | 0.659 - 0.670 |
| Vendor 2 | 3D-res | 0.31 - 10.88 | 14 | 0.674 | 0.669 - 0.677 |
| Vendor 2 | 3D-res | 0.31 - 10.88 | 15 | 0.668 | 0.662 - 0.671 |

### 5. AI models trained on phantom-only and patient-only data

#### 5.1 Lesion characteristics of training data

**Supplementary Fig. 7.**
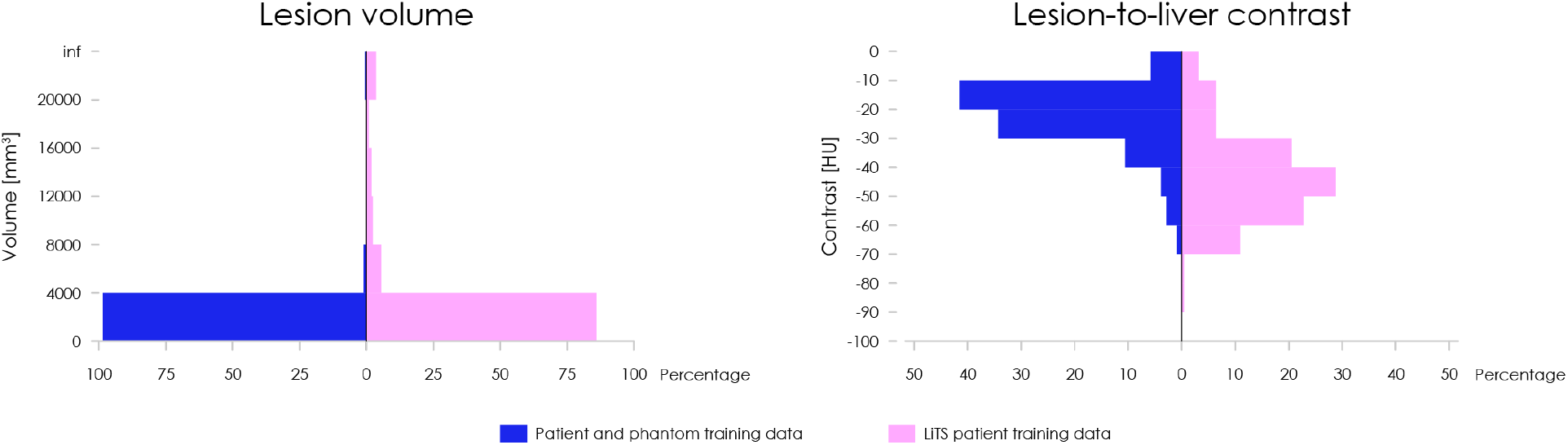
Lesion characteristics of training datasets. Distribution of lesion volumes and lesion-to-liver contrast for the phantom-only and corresponding patient-only training data (blue) and the Liver Tumor Segmentation (LiTS) patient training data (pink).

#### 5.2 Clinical in-house test cohort

**Supplementary Table 4.** Characteristics of the in-house clinical test cohort. CT systems and volume computed tomography indices (CTDIvol) of the retrospectively collected in-house test data.

| CT system | Number of cases | CTDIvol range (mGy) |
| --- | --- | --- |
| Canon Aquilion PRIME | 15 | 3.2 – 10.2 |
| Canon Aquilion One | 7 | 1.6 – 5.1 |
| Canon Aquilion Lightning | 1 | 7.7 |
| GE Revolution HD | 6 | 5.89 – 13.73 |
| GE Revolution EVO | 6 | 6.82 – 10.88 |
| GE Revolution CT | 5 | 6.32 – 9.19 |
| Siemens SOMATOM go.top | 1 | 7.0 |
| Siemens Perspective | 1 | 3.4 |
| Siemens Emotion16 | 1 | 7.4 |

#### 5.3 Recall and precision

**Supplementary Fig. 8.**
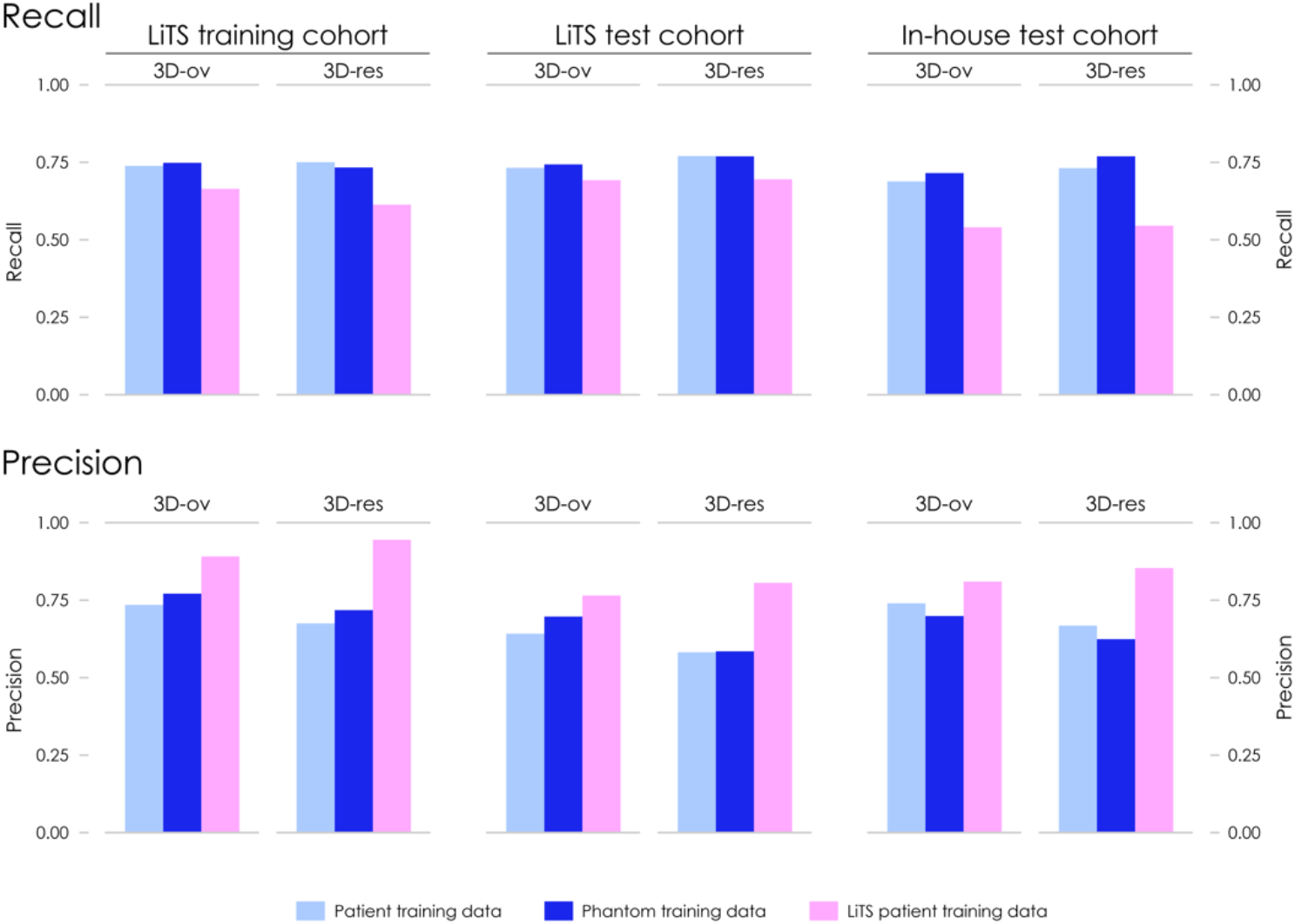
Recall and precision of AI models trained on phantom-only and patient-only data. Results are shown for both nnU-Net architectures, the original 3D full-resolution configuration (3D-ov) and the modified variant with residual encoder connections (3D-res), across three clinical test cohorts: the Liver Tumor Segmentation (LiTS) training cohort excluding the five cases included in the LiTS subcohort training set, the LiTS test cohort, and an in-house test cohort.

#### 5.4 Numerical results

**Supplementary Table 5.** F1 scores, recall and precision of AI models trained on phantom-only and patient-only data. Results are presented for both nnU-Net architectures, the original 3D full-resolution configuration (3D-ov) and the modified variant with residual encoder connections (3D-res), across three clinical test cohorts: the Liver Tumor Segmentation (LiTS) training cohort excluding the five cases included in the LiTS subcohort training set, the LiTS test cohort, and an in-house test cohort.

| nnU-Net architecture | Training data | Test data | F1 score | Recall | Precision |
| --- | --- | --- | --- | --- | --- |
| 3D-ov | Patient cohort | In-house cohort | 0.663 | 0.652 | 0.673 |
| 3D-ov | Phantom cohort | In-house cohort | 0.659 | 0.647 | 0.671 |
| 3D-ov | LiTS subcohort | In-house cohort | 0.65 | 0.551 | 0.791 |
| 3D-ov | Patient cohort | LiTS training cohort | 0.703 | 0.76 | 0.655 |
| 3D-ov | Phantom cohort | LiTS training cohort | 0.731 | 0.758 | 0.705 |
| 3D-ov | LiTS subcohort | LiTS training cohort | 0.729 | 0.69 | 0.772 |
| 3D-ov | Patient cohort | LiTS test cohort | 0.733 | 0.731 | 0.735 |
| 3D-ov | Phantom cohort | LiTS test cohort | 0.756 | 0.742 | 0.771 |
| 3D-ov | LiTS subcohort | LiTS test cohort | 0.778 | 0.691 | 0.891 |
| 3D-res | Patient cohort | In-house cohort | 0.653 | 0.71 | 0.605 |
| 3D-res | Phantom cohort | In-house cohort | 0.655 | 0.719 | 0.601 |
| 3D-res | LiTS subcohort | In-house cohort | 0.644 | 0.524 | 0.836 |
| 3D-res | Patient cohort | LiTS training cohort | 0.673 | 0.77 | 0.598 |
| 3D-res | Phantom cohort | LiTS training cohort | 0.66 | 0.736 | 0.598 |
| 3D-res | LiTS subcohort | LiTS training cohort | 0.713 | 0.637 | 0.81 |
| 3D-res | Patient cohort | LiTS test cohort | 0.719 | 0.769 | 0.675 |
| 3D-res | Phantom cohort | LiTS test cohort | 0.742 | 0.768 | 0.718 |
| 3D-res | LiTS subcohort | LiTS test cohort | 0.8 | 0.694 | 0.945 |

### 6. AI models trained on phantom-data-augmented datasets

#### 6.1 Recall and precision in phantom test data

**Supplementary Fig. 9.**
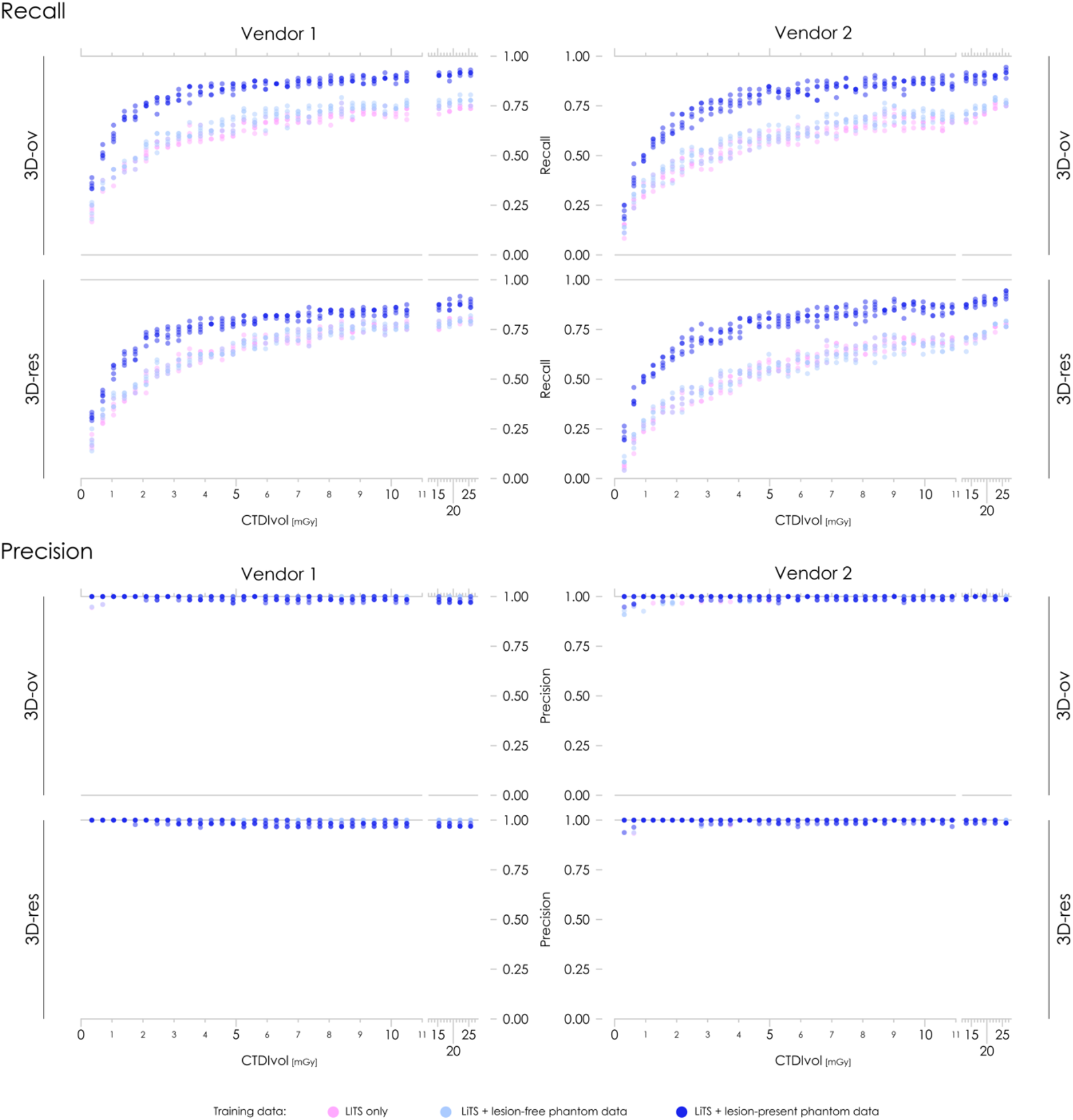
Recall and precision in phantom test data following phantom data-augmented AI training. Results are shown for both nnU-Net architectures, the original 3D full-resolution configuration (3D-ov) and the modified variant with residual encoder connections (3D-res), both scanner systems, all dose levels, and five repeated acquisitions. CTDIvol is the volume computed tomography dose index.

#### 6.2 Recall and precision in patient test data

**Supplementary Fig. 10.**
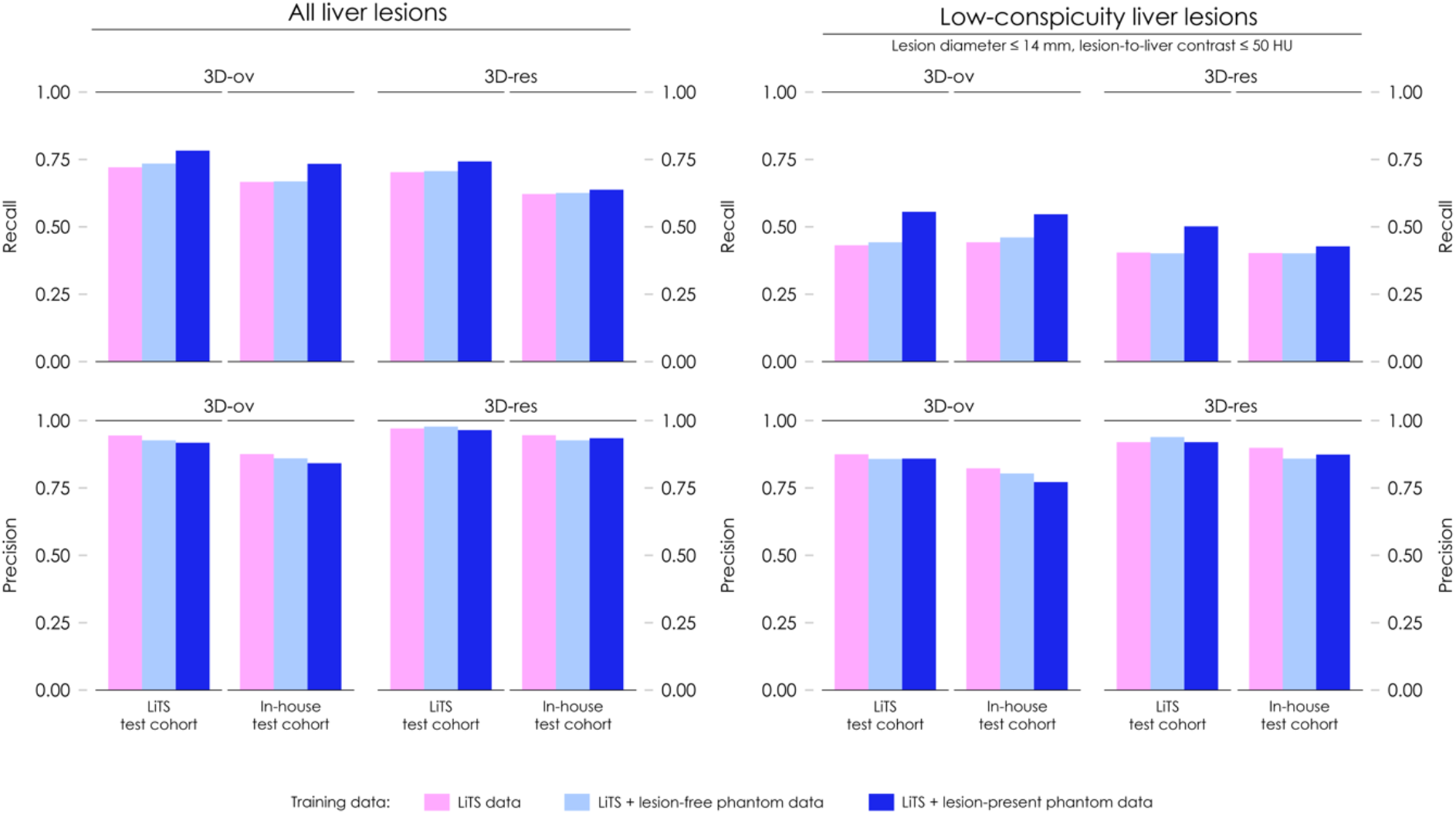
Recall and precision in clinical test data following phantom data-augmented AI training. Results are shown for both nnU-Net architectures, the original 3D full-resolution configuration (3D-ov) and the modified variant with residual encoder connections (3D-res), across two clinical test cohorts: the Liver Tumor Segmentation (LiTS) test cohort and an in-house test cohort. The subanalysis for low-conspicuity lesions includes lesions 14 mm or smaller in diameter and -50 HU or greater lesion-to-liver contrast.

#### 6.3 Recall and precision by lesion subgroup in phantom test data

**Supplementary Fig. 11.**
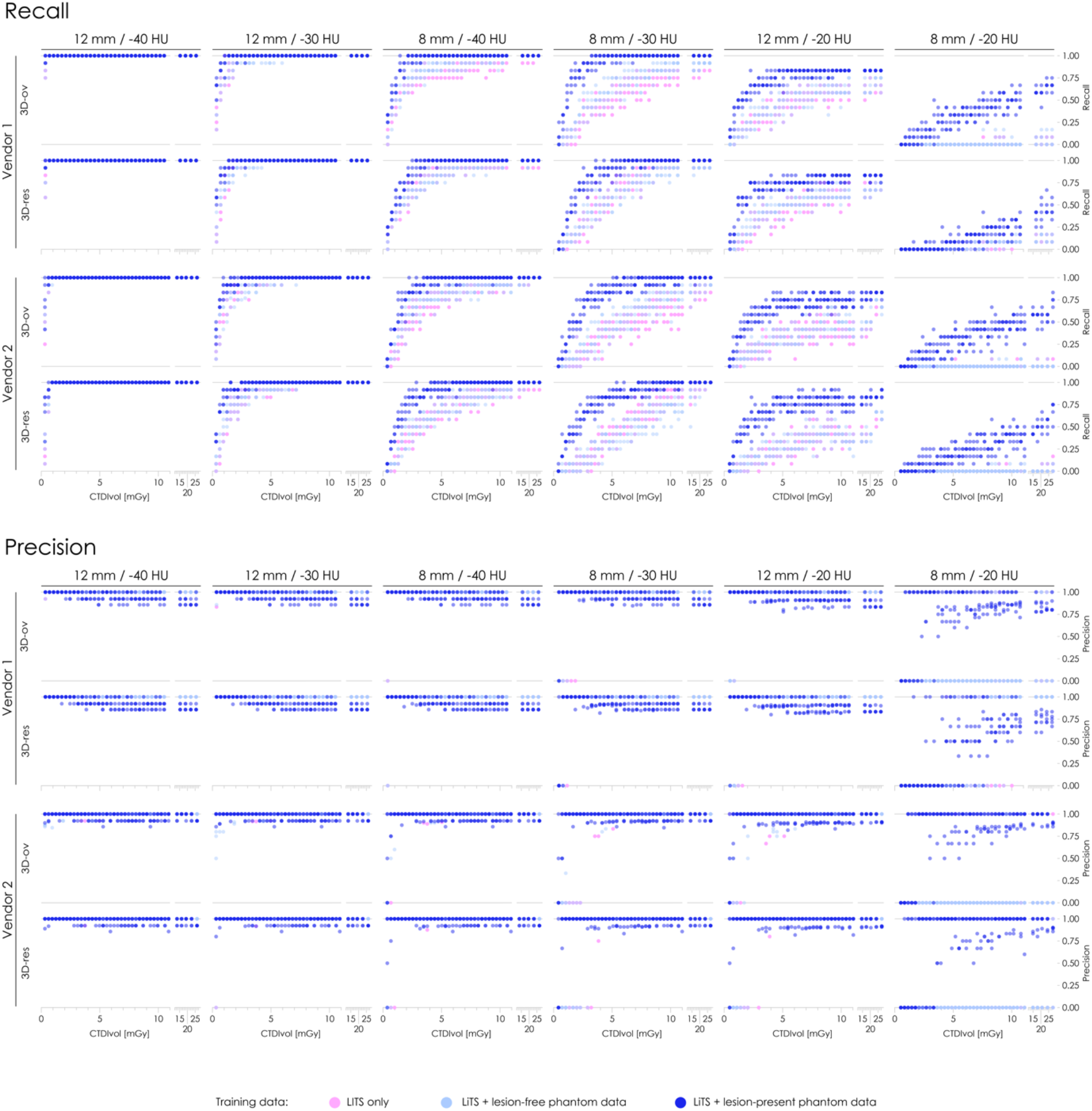
Recall and precision following phantom data-augmented AI training across liver lesion subgroups in phantom test data. Results are presented for for each lesion diameter and lesion-to-liver contrast combination labelled at the top of each panel across both scanner systems and nnU-Net architectures: the original 3D full-resolution configuration (3D-ov) and the modified variant with residual encoder connections (3D-res). CTDIvol is the volume computed tomography dose index.

## References

1. Rajpurkar, P. & Lungren, M.P. The Current and Future State of AI Interpretation of Medical Images. N Engl J Med 388, 1981–1990 (2023).

2. Brady, A.P., et al. Developing, Purchasing, Implementing and Monitoring AI Tools in Radiology: Practical Considerations. A Multi-Society Statement from the ACR, CAR, ESR, RANZCR and RSNA. Radiol Artif Intell 6, e230513 (2024).

3. Kilim, O., et al. Physical imaging parameter variation drives domain shift. Sci Rep 12, 21302 (2022).

4. Blazis, S.P., Dickerscheid, D.B.M., Linsen, P.V.M. & Martins Jarnalo, C.O. Effect of CT reconstruction settings on the performance of a deep learning based lung nodule CAD system. Eur J Radiol 136, 109526 (2021).

5. Yu, A.C., Mohajer, B. & Eng, J. External Validation of Deep Learning Algorithms for Radiologic Diagnosis: A Systematic Review. Radiology: Artificial Intelligence 4, e210064 (2022).

6. Allen, B., et al. Evaluation and Real-World Performance Monitoring of Artificial Intelligence Models in Clinical Practice: Try It, Buy It, Check It. J Am Coll Radiol 18, 1489–1496 (2021).

7. de Vries, C.F., et al. Impact of Different Mammography Systems on Artificial Intelligence Performance in Breast Cancer Screening. Radiol Artif Intell 5, e220146 (2023).

8. American College of Radiology and Society for Imaging Informatics in Medicine. ACR-SIIM Practice Parameter for Imaging Artificial Intelligence (AI). (Washington, DC, 2026).

9. Coombs, L.P., et al. ACR’s Assess-AI: A Registry for Real-World Performance Monitoring of Clinical Imaging Artificial Intelligence. J Am Coll Radiol (2026).

10. Feng, J., et al. Clinical artificial intelligence quality improvement: towards continual monitoring and updating of AI algorithms in healthcare. NPJ Digit Med 5, 66 (2022).

11. Judy, P.F., et al. Phantoms for Performance Evaluation and Quality Assurance of CT Scanners. (Alexandria, VA, 1977).

12. Bellon, E.M., Miraldi, F.D. & Wiesen, E.J. Performance of evaluation of computed tomography scanners using a phantom model. AJR Am J Roentgenol 132, 345–352 (1979).

13. Conzelmann, J., Schwarz, F.B., Hamm, B., Scheel, M. & Jahnke, P. Development of a method to create uniform phantoms for task-based assessment of CT image quality. J Appl Clin Med Phys 21, 201–208 (2020).

14. Ardila Pardo, G.L., et al. 3D printing of anatomically realistic phantoms with detection tasks to assess the diagnostic performance of CT images. Eur Radiol 30, 4557–4563 (2020).

15. Vaishnav, J.Y., Jung, W.C., Popescu, L.M., Zeng, R. & Myers, K.J. Objective assessment of image quality and dose reduction in CT iterative reconstruction. Med Phys 41, 071904 (2014).

16. Bellmann, Q., et al. Low-contrast lesion detection in neck CT: a multireader study comparing deep learning, iterative, and filtered back projection reconstructions using realistic phantoms. Eur Radiol Exp 8, 84 (2024).

17. Jahnke, P., et al. Radiopaque Three-dimensional Printing: A Method to Create Realistic CT Phantoms. Radiology 282, 569–575 (2017).

18. Jahnke, P., et al. Paper-based 3D printing of anthropomorphic CT phantoms: Feasibility of two construction techniques. Eur Radiol 29, 1384–1390 (2019).

19. Mei, K., et al. Three-dimensional printing of patient-specific lung phantoms for CT imaging: Emulating lung tissue with accurate attenuation profiles and textures. Med Phys 49, 825–835 (2022).

20. Schikorra, T., et al. Validation of a novel 3D-printed anthropomorphic pediatric abdomen phantom using photon-counting CT. Sci Rep 15, 43812 (2025).

21. Abrams, H.L., Spiro, R. & Goldstein, N. Metastases in carcinoma; analysis of 1000 autopsied cases. Cancer 3, 74–85 (1950).

22. Bosch, F.X., Ribes, J. & Borras, J. Epidemiology of primary liver cancer. Semin Liver Dis 19, 271–285 (1999).

23. Isensee, F., Jaeger, P.F., Kohl, S.A.A., Petersen, J. & Maier-Hein, K.H. nnU-Net: a self-configuring method for deep learning-based biomedical image segmentation. Nat Methods 18, 203–211 (2021).

24. Isensee, F., et al. nnU-Net Revisited: A Call for Rigorous Validation in 3D Medical Image Segmentation. in Medical Image Computing and Computer Assisted Intervention – MICCAI 2024 (eds. Linguraru, M.G., et al.) 488–498 (Springer Nature Switzerland, Cham, 2024).

25. Bilic, P., et al. The Liver Tumor Segmentation Benchmark (LiTS), (2019).

26. Huang, K., et al. Impact of slice thickness, pixel size, and CT dose on the performance of automatic contouring algorithms. J Appl Clin Med Phys 22, 168–174 (2021).

27. Sandfort, V., Yan, K., Pickhardt, P.J. & Summers, R.M. Data augmentation using generative adversarial networks (CycleGAN) to improve generalizability in CT segmentation tasks. Sci Rep 9, 16884 (2019).

28. Koetzier, L.R., et al. Generating Synthetic Data for Medical Imaging. Radiology 312, e232471 (2024).

29. Larson, D.B., Bhargavan-Chatfield, M., Tilkin, M., Coombs, L. & Wald, C. The Road Map for ACR Practice Accreditation for Radiology Artificial Intelligence. J Am Coll Radiol 22, 586–592 (2025).

30. Wang, G., et al. Development of metaverse for intelligent healthcare. Nat Mach Intell 4, 922–929 (2022).

31. Samei, E., et al. Performance evaluation of computed tomography systems: Summary of AAPM Task Group 233. Med Phys 46, e735–e756 (2019).

32. Yan, L., et al. Size and Contrast Thresholds for Liver Lesion Detection in Regular and Low-dose CT Examinations: A Reader Study of 2300 Synthetic Lesions Across 100 Patients. Acad Radiol 32, 3952–3959 (2025).

33. Michallek, F., Genske, U., Niehues, S.M., Hamm, B. & Jahnke, P. Deep learning reconstruction improves radiomics feature stability and discriminative power in abdominal CT imaging: a phantom study. Eur Radiol 32, 4587–4595 (2022).

34. Martin Asiain, M., et al. Impact of CT dose on AI performance: A comparison of radiomics, deep, and foundation models in a multicentric anthropomorphic phantom study. Med Phys 53, e70374 (2026).

35. Goelz, L., et al. Inconsistency of AI in intracranial aneurysm detection with varying dose and image reconstruction. Sci Rep 15, 19921 (2025).

36. Alstrom, L., et al. Assessing the robustness of AI lesion risk scores at different exposure settings using an anthropomorphic breast phantom. Radiat Prot Dosimetry 202, 220–228 (2026).

37. Racine, D., et al. Task-based quantification of image quality using a model observer in abdominal CT: a multicentre study. Eur Radiol 28, 5203–5210 (2018).

38. Roth, H., et al. A new 2.5 D representation for lymph node detection in CT. (The Cancer Imaging Archive, 2015).

39. Simpson, A.L., et al. A large annotated medical image dataset for the development and evaluation of segmentation algorithms. arXiv (2019).

